# EAGLE-AI: A large language model workflow for automated extraction and scoring of literature evidence linking genes to autism spectrum disorder

**DOI:** 10.1101/2025.09.10.25334730

**Authors:** Vinicius Furlan, Julian Moran, Nelson B. Salazar, Olivia Rennie, Ny Hoang, Andrew Wan, Marla Mendes de Aquino, Worrawat Engchuan, Jacob A.S. Vorstman, Stephen W. Scherer

## Abstract

We previously developed the Evaluation of Autism Gene Link Evidence (EAGLE) curation framework and usit to characterise 219 autism-associated genes. However, this took years of human work. We present EAGLE-AI, an automated curation system incorporating large language models (LLMs). On screened paper sets, it achieves F1 91% and scoring error 17.2%, near human performance. EAGLE-AI performs worse on unscreened papers due to table parsing and context overload, which we address using if-else scoring and computer vision tools. Handling of supplementary materials remains an unsolved problem. Our findings demonstrate proof-of-concept for automating most of a clinical genomics curation process.

## Background

Next-generation sequencing (NGS) has greatly improved our ability to detect genetic variants across the human genome^1^. In leveraging NGS and related genetic testing methods, laboratories are achieving a greater diagnostic yield for many disorders, including autism spectrum disorder (ASD, also called autism)^2^. However, this relies on enough scientific understanding to determine the significance of the genetic variants identified. Here, knowing whether a gene is related to a condition is a crucial first step^3^. In the case of ASD, research has demonstrated a significant genetic component driving the condition^4,5,6,7,8^.

Previous studies have shown considerable inconsistency in the genes included in autism gene panels^9,10^. This highlights the need to distinguish between bona-fide confirmed genes and candidate genes, referred to as establishing gene-phenotype clinical validity^11^. To promote consistency among laboratories, researchers, and clinicians, the Clinical Genome Resource (ClinGen) has developed a framework for systematically evaluating the evidence supporting a gene-phenotype association^12^. This framework involves curating publicly available evidence, including genetic and experimental data, and applying a rules-based scoring system. The evidence level is categorized as definitive, strong, moderate, or limited based on the strength of evidence. With this framework, teams of hundreds of ClinGen volunteers from the United States and around the world have evaluated more than 2,420 gene-phenotype relationships and established several gene curation expert panels, including one for Intellectual Disability and Autism^13,14,15^.

Intellectual disability and ASD are often grouped under the umbrella term of neurodevelopmental disorders (NDDs). As an indication for genetic testing and clinical consultation, NDD as a category is commonly accepted by clinical laboratories and genetic clinics, which is reasonable given the high comorbidity among the conditions included in this general term^11,16^. However, for gene curation purposes, it is important to assess the clinical validity for ASD independently from NDDs as a whole, as the implications in clinical management may differ depending on the gene annotation. For instance, ASD often manifests independently of intellectual disability, epilepsy, or other NDDs^17,18^. Moreover, the support needs differ between individuals with ASD and those with both ASD and other co-occurring NDDs^19,20^. As genome sequencing is incorporated into clinical care more widely and at earlier stages of development, understanding a gene’s relevance to ASD specifically will better empower precision care^21^. Efforts to parse the heterogeneity of autism are ongoing: for example, recent work has identified clinically distinct autism subtypes linked to specific genetic programs and developmental trajectories^22,23,24,25^.

To this end, we previously established the Evaluation of Autism Gene Link Evidence (EAGLE) framework for curating ASD clinical genomics evidence^10^. This approach builds on the ClinGen gene-phenotype clinical validity framework by adding a rules-based method for assessing level of confidence in the reported ASD phenotype. EAGLE also adjusts the genetic evidence scoring approach to account for the variability and complexity of ASD genetics. EAGLE reserves high scores for variants linked to a confirmed ASD diagnosis rather than a case with autism-like features. This helps in distinguishing ASD-associated variants from variants linked to NDDs more broadly.

Previously, ASD gene lists have been defined from large-scale sequencing studies of individuals with ASD. They include lists from the Simons Foundation Powering Autism Research for Knowledge (SPARK)^26^, Autism Speaks – MSSNG^27^, and the Autism Sequencing Consortium (ASC)^28^. Together, these three projects identify a total of 236 unique genes as being associated with ASD with high confidence. In contrast, EAGLE is used to define an autism gene list based on evidence from the full diversity of the scientific literature^10^.

EAGLE scores are currently included in the Simons Foundation Autism Research Initiative (SFARI) database^29^ and characterize level of evidence for many prototypical ASD genes: for example, *NRXN1*, *SCN2A*, *MECP2*, *CHD8, DDX3X, SHANK3*, and *PTEN*. However, the EAGLE framework’s manual curation workflow is both time-consuming and resource-intensive, as it requires in-depth assessment of ASD clinical case reports, functional studies, and database information.

Since at least 2003, research groups have been using natural language processing models to automate or semi-automate literature curation steps scalably^30^. Transformer architectures ushered in a new wave of interest: for instance, models based on Bidirectional Encoder Representations from Transformers (BERT) have been used to automate literature data extraction and paper classification tasks^31,32^. During the COVID-19 pandemic, a variety of automation resources emerged for dealing with the onslaught of actionable scientific literature^32,33,34^. With the more recent advent of large language models (LLMs) such as Generative Pre-trained Transformers (GPTs), other research groups have leveraged these models in automating literature review, such as for paper screening or data extraction^24,25,26^. However, most peer-reviewed work in this area has focused on automating only a single workflow phase^35^.

In a similar vein, and to address the costs of manual literature curation in the context of ASD clinical genomics, we developed EAGLE-AI, an evidence collection, screening, extraction, and scoring workflow that incorporates LLMs. This workflow aims to accelerate the accurate identification of ASD-associated genes by operating, with minimal manual intervention, on a large and evolving corpus of literature (Fig. 1).

**Fig. 1.**
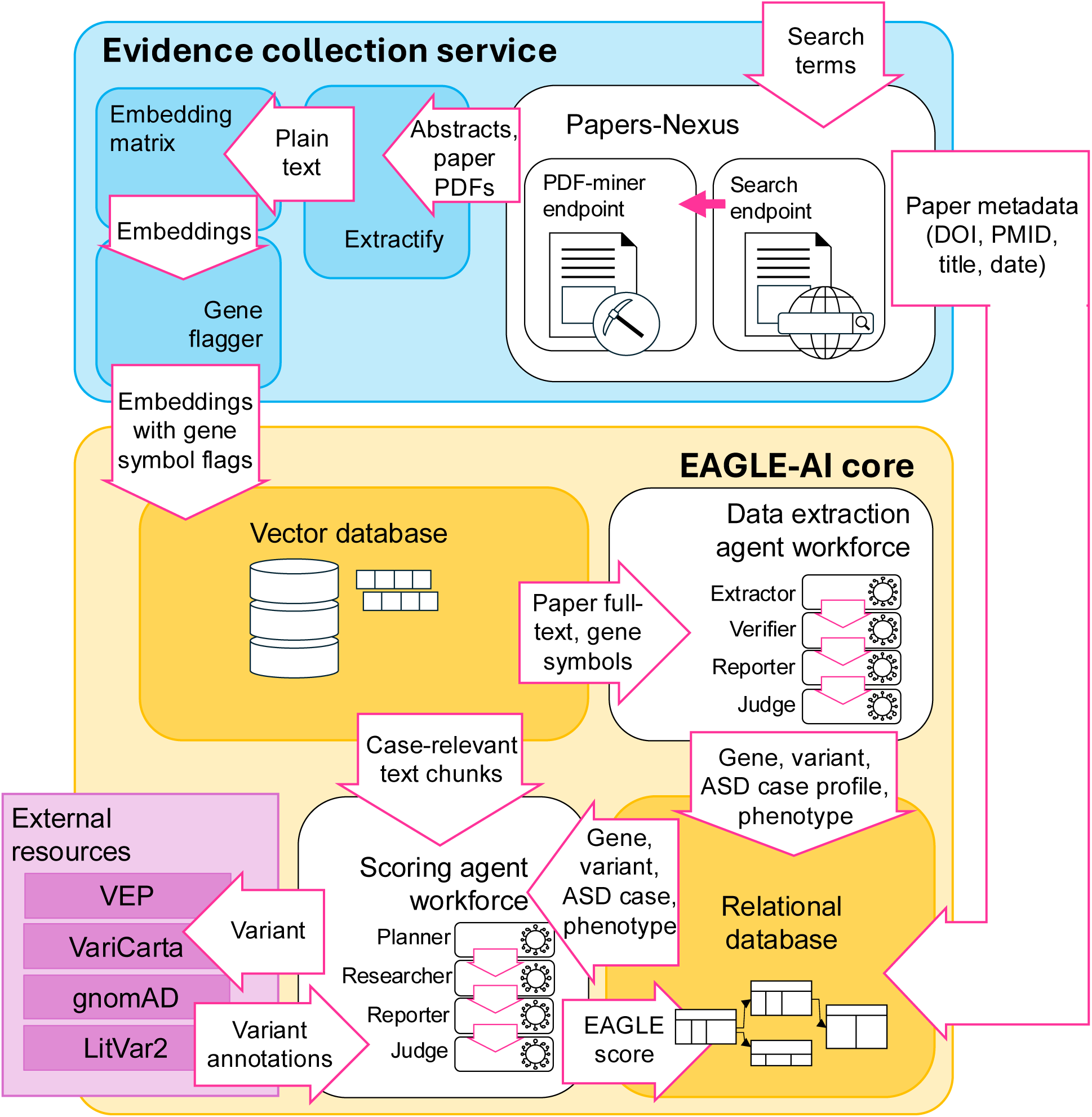
EAGLE-AI system architecture. Schematic of EAGLE-AI’s whole-system architecture, with initial input consisting of natural-language search terms and final output consisting of the populated relational database. Search terms consist of a gene name (e.g. “*DMD*”) and the fixed search terms “ASD” and “autism”. Database entries are dedicated to single ASD cases and include a case patient description, phenotype description, gene name, genomic variant ID, and evidence score.

The data extraction and scoring modules of EAGLE-AI both deploy multi-agent workforces using o3-mini^36^ and GPT-4o^37,38^ (Fig. 2). We developed two additional versions of EAGLE-AI: one incorporating a BiLSTM-CRF model, trained in-house for named entity recognition (NER), and another deploying a Llama 3.1-8B model finetuned for EAGLE-compliant data extraction.

**Fig. 2.**
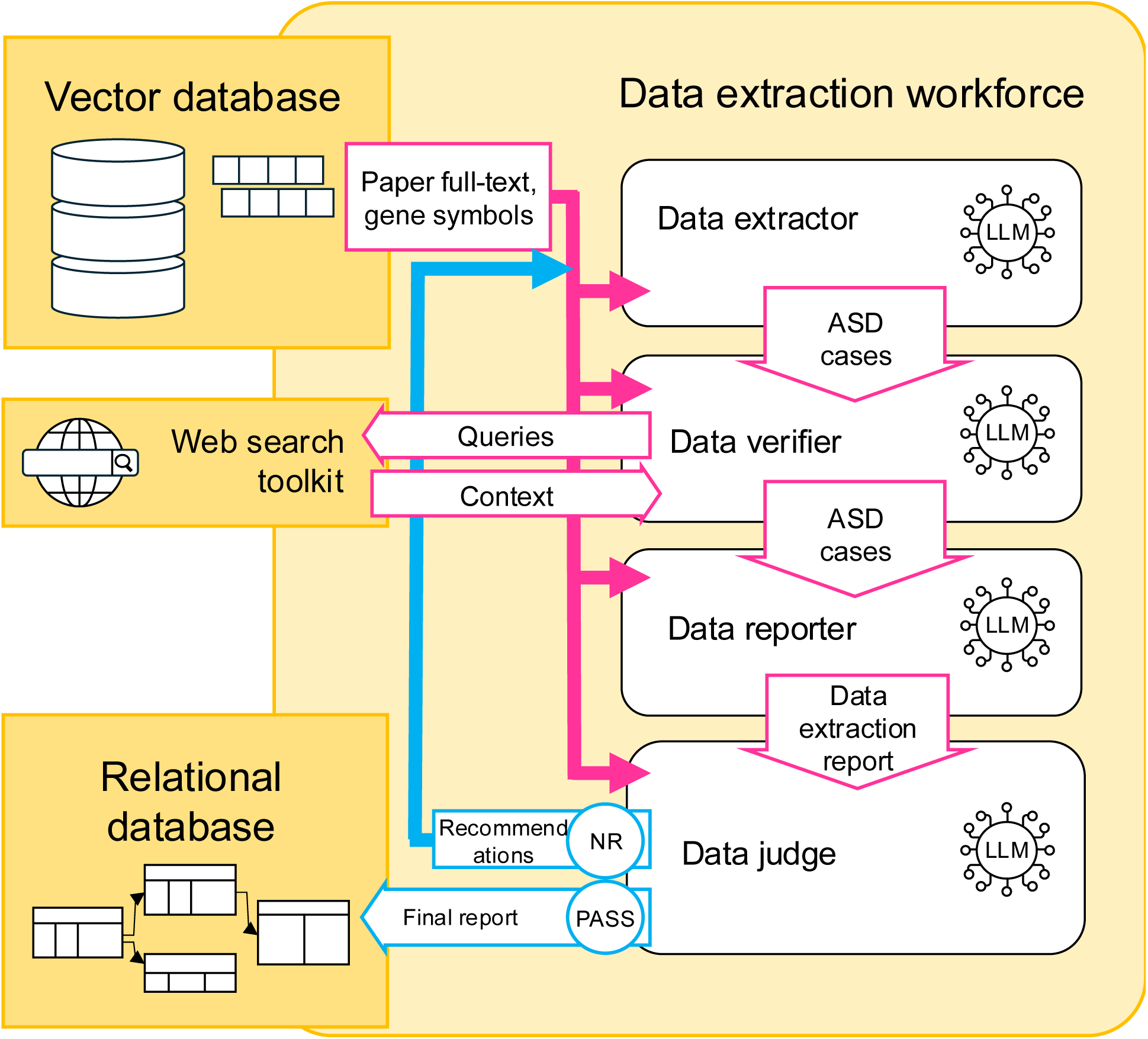
Agentic EAGLE-AI core data extraction workflow. Schematic of Eagle-AI core’s agentic data extraction workforce. The workforce includes an additional task planner and coordinator agent for incorporating recommendations from the judge in the event of a “NEEDS_REVISION” (NR) verdict. Depending on the recommendations, this can yield revision tasks to any of the four workforce agents. The final output report complies with the format presented in Table S1.

EAGLE-AI showcases not only the power of third-party LLMs in extracting data from literature, but also their lesser suitability for evidence scoring when a deterministic rules-based algorithm can be applied instead. We expect that EAGLE-AI’s system architecture can generalize, in modular fashion, to automate important aspects of evidence curation for other clinical genomics research questions.

## Results

### System workflow

In evaluating the association between a genomic variant and ASD, the manual EAGLE curation standard assesses evidence from three categories: genetic evidence, quality of ASD diagnostic evaluation, and experimental evidence (e.g. animal models) (Table 1). Because the EAGLE curation standard is rigorously rules-based, EAGLE-AI implements the same rules in automated fashion for the tasks of data extraction and scoring. As such, the system is comprised of two interconnected modules: an evidence collection service and EAGLE-AI core, which performs the data extraction and scoring steps (Fig. 1).

The evidence collection service maintains a list of genes currently under investigation and submits requests to Papers-Nexus, a purpose-built web API for automatically retrieving and downloading open-access full-text PDFs from PubMed, Elsevier, and Google Scholar using search terms. To keep the EAGLE-AI databases up to date, the evidence collection service requests papers pertaining to a given gene on a scheduled periodic basis.

The evidence collection service subsequently processes the collected papers by converting the PDFs to plain text, extracting table contents, and composing image-to-text descriptions of the paper figures. All text extracted from the papers is converted to embeddings using OpenAI’s text-embedding-3-large matrix and stored in a vector database.

The EAGLE-AI workflow handles internal data storage using two distinct databases (Fig. 1). It stores tabular data containing string entries, i.e. paper metadata, extracted cases, and variant information, in a relational database. Embeddings representing each paper’s text are indexed using a vector database. Using an API endpoint for o3-mini^36^, EAGLE-AI assigns semantic flags to each text chunk, where the flags pertain to genes, phenotype terms, and variants.

EAGLE-AI core deploys eight distinct LLM agents, four of which are dedicated to data extraction and four of which are dedicated to evidence-scoring each gene and candidate ASD case (Fig. 1). The data extraction agents write to the relational database, using a tabular format populated with detailed information on the ASD case subject, gene, etiological genomic variant, and phenotype (Table S1). EAGLE-AI core’s scoring agents add a final score, indicating the strength of ASD etiological evidence, to the relational database as well. The relational database is therefore the final repository of EAGLE-AI’s data extractions and evidence scores.

### Evidence collection service workflow

EAGLE-AI’s evidence collection service submits requests to Papers-Nexus, an API developed in-house for performing automated English-language literature searches and bulk full-text retrieval from search queries. To find relevant paper metadata, EAGLE-AI submits automated requests to the search endpoint of Papers-Nexus, where the request includes a general search term field, a gene search term field, and an exclusion term field. Search criteria consist of a gene name (e.g. “*DMD*”) and the fixed search terms “ASD” and “autism”.

Using the Bio.Entrez^39^ and scholarly^40^ Python packages, Papers-Nexus searches for relevant papers in the PubMed, Google Scholar, and Elsevier databases. Its response to the client provides the paper title, author name(s), abstract, PMID, DOI, and citation information for each paper that satisfies the search criteria.

In light of our design goal to have EAGLE-AI extract data from a large corpus of English-language papers with minimal manual input, we developed a PDF-miner endpoint for Papers-Nexus. This endpoint is dedicated to bulk downloading open-access full-text PDFs of papers, where the client’s request contains a list of PMIDs or DOIs. Papers-Nexus retrieves full-text PDFs using the PubMed-OA FTP server^41^ and the Unpaywall API^42^.

Extractify, a module within EAGLE-AI’s evidence collection service, uses Adobe’s pdf.services^43^ and openpyxl^44^ to convert retrieved papers and supplementary tables to plain text. Paper tables are converted to markdown using GPT-4o^37^. For each paper, a gene flagger module deploys an o3-mini model to extract HGNC symbols for all ASD-relevant genes mentioned by the paper, as well as paper metadata. The gene symbols are assigned as flags to the paper’s full text. Here, gene symbols are flagged only if the paper presents clinical or experimental evidence linking the gene to ASD.

### Evidence collection service retrieval rate

Using a test set of metadata for 172 papers from the PubMed database, we found that Papers-Nexus retrieved full-text PDFs for 114 (66.3%) of the papers. For all remaining papers, Papers-Nexus retrieved the title, author names, and abstract. Papers-Nexus failed to retrieve a paper’s full-text if the paper is not open-access or if the paper’s host denies transparent requests to automatically download the PDF.

### Agentic data extraction workflow

EAGLE-AI’s data extraction workforce is responsible for extracting three categories of relevant information required for scoring: genomic variant, case ID, and phenotype. It deploys four distinct LLM agents, which perform automated tasks based on their own decision-making from inputs. The agents are assigned to the roles of data extractor, data verifier, data reporter, and data extraction judge. As of the 2025 version of EAGLE-AI core, the data extractor, verifier, and judge use OpenAI’s o3-mini model^36^, whereas the reporter uses GPT-4o^37^. The agentic workforce is managed using the CAMEL-AI multi-agent framework^45^ Input to the data extraction workforce consists of a single paper’s plain full-text and all gene symbols flagged by the gene flagger (Fig. 2).

For each gene in the paper, the data extractor agent receives a prompt that includes the given gene symbol, instructing the extractor to identify and extract all ASD cases in the paper linked to the given gene. For each case, the case subject must have an ASD diagnosis under specified criteria; the gene must be described with identifying detail; and the paper must describe a specific ASD-driving genomic variant. For all eligible cases, the extractor gives as output the case subject’s identifier, the location of the text where the case is described, a brief description of the variant, and an account of pertinent data missing from the paper (Table S2).

A prompt to the verifier agent instructs it to review all sections of the paper full-text and identify any cases missed by the extractor. Using CAMEL’s SearchToolkit, the verifier is able seek further information through web search^46^. For each case, its output format resembles that of the extractor (Table S3).

The reporter agent’s task is to synthesize all cases from the extractor and verifier. The reporter is instructed to remove any duplicate cases and to reformat the data to match the example presented in Table S1, which in turn matches the schema of EAGLE-AI’s relational database.

The judge agent assesses the reporter’s output for quality and consistency. In evaluating quality, the judge checks for whether the data types of populated fields comply with the database schema and whether gene and variant details are logically consistent. The judge flags cases with insufficient details about the gene or candidate ASD-driving variant. It also flags any potentially duplicate cases. Its output includes an overall quality score of high, medium, or low (Table S4). Additionally, it issues a verdict of the gene data from the paper: namely, as “PASS” or “NEEDS_REVISION”.

As per the CAMEL-AI multi-agent framework^45^, an additional task planner agent and coordinator agent handle task assignment to the workforce beyond the initial prompts. In cases where the judge issues a verdict of “NEEDS_REVISION”, these agents assign new tasks to the extractor, verifier, reporter, and/or judge based on the judge’s account of the issues. The workforce’s passed output writes to EAGLE-AI core’s relational database.

### EAGLE-AI data extraction precision-recall

We evaluated the precision-recall of EAGLE-AI core’s data extraction module while using four different configurations (Fig. 3) as described in Methods. Here, we measured how well the data extraction module reproduced the extractions of the EAGLE manual curators. We evaluated all four configurations of EAGLE-AI core on the same set of 116 publications and 96 cases screened for ease of machine-readability (see Methods for screening criteria). All cases were reported as having a variant in *DMD, SHANK1*, *CACNA1D*, *DDX3X*, *DDX53*, or *MECP2*. Raw performance metrics are included in Table S5, Table S6, and Table S7. Square brackets give the 95% confidence intervals.

**Fig. 3.**
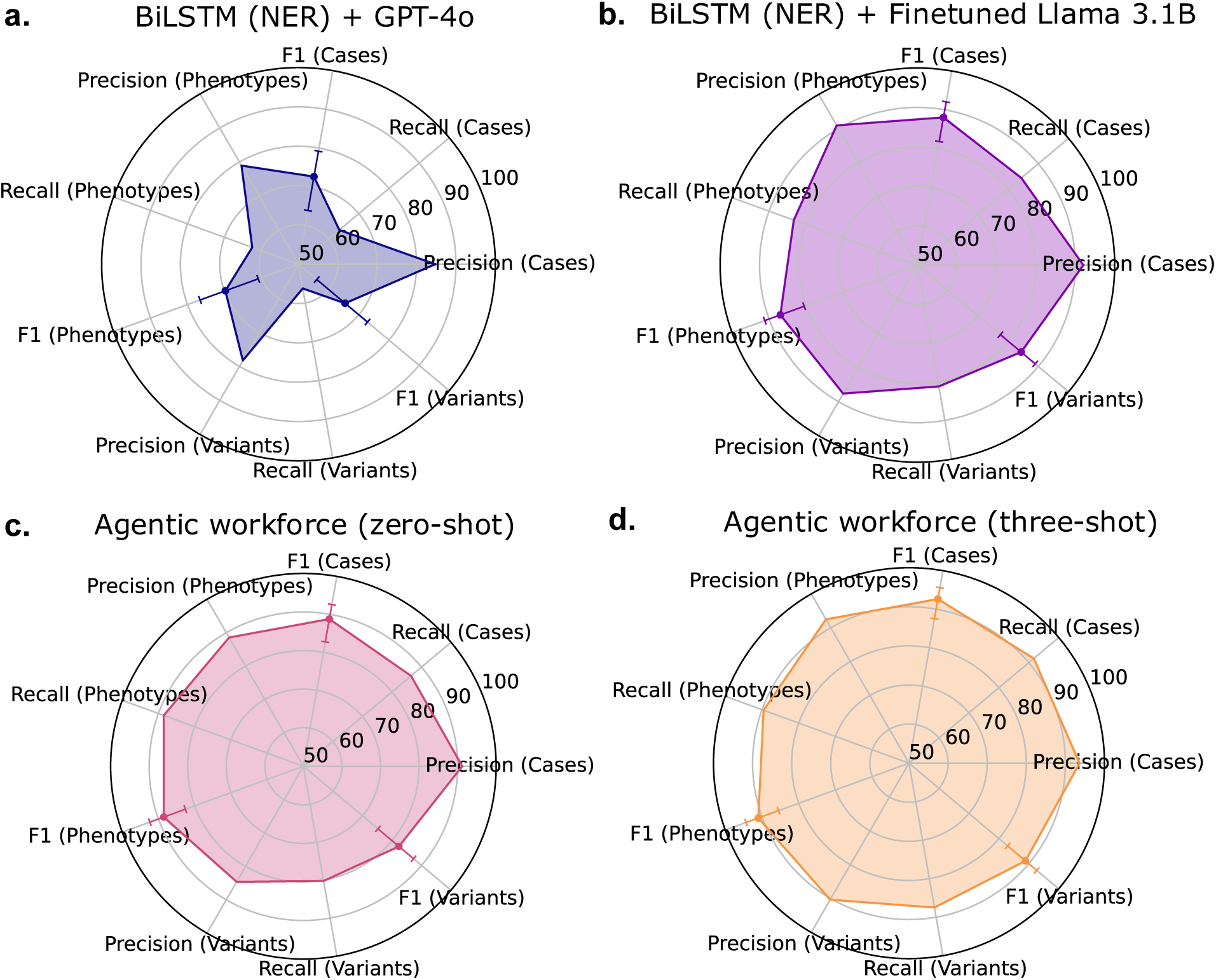
EAGLE-AI data extraction performance. EAGLE-AI’s data extraction precision-recall metrics out of 100 using four different approaches, evaluated across manually scored cases from 116 papers about *DMD, SHANK1, CACNA1D, DDX3X, DDX53*, or *MECP2*; error bars indicate 95% F1 confidence intervals. **a** Extraction performance of the NER model for relevance filtering and text chunk assembly, coupled with a GPT-4o endpoint for data extraction. **b** Extraction performance of the NER model coupled with a finetuned Llama 3.1-8B model for data extraction. **c** Extraction performance of agentic EAGLE-AI core using a zero-shot task prompt to the data extraction workforce. **d** Extraction performance of agentic EAGLE-AI core using a three-shot task prompt to the data extraction workforce.

The NER model achieved an average F1 of 69%, with case F1 of 73% [64%, 79%], variant F1 of 65% [56%, 73%], and phenotype F1 of 70% [61%, 77%] (Fig. 3a).

Finetuned Llama 3.1-8B achieved average F1 of 86%, with case F1 of 88% [82%, 92%], variant F1 of 84% [77%, 89%], and phenotype F1 of 87% [81%, 91%] (Fig. 3b).

The zero-shot agentic workforce achieved an average F1 score of 86%, with case F1 of 89% [83%, 93%], variant F1 of 82% [75%, 87%], and phenotype F1 of 89% [83%, 92%] (Fig. 3c).

The three-shot agentic workforce achieved average F1 of 91%, with case F1 of 93% [88%, 96%], variant F1 of 89% [83%, 93%], and phenotype F1 of 91% [86%, 94%] (Fig. 3d).

Table S8 presents an example whereby, after finetuning, the Llama 3.1-*B model was better able to identify the pathogenicity impact of a *DDX3X* variant NM_001356.5:c.1171-2A>G as reported in a paper abstract and case patient description. For a case with a *MECP2* indel variant, Llama 3.1-8B assigned the accurate phenotype confidence level (low) only after finetuning, resulting in an EAGLE score closer to the manual ground truth (Table S9).

### Agentic evidence scoring workflow

Fig. 4 presents a schematic of EAGLE-AI core’s multi-agent workforce for scoring possible ASD cases and their reported variants. Much like the data extraction workforce, the scoring workforce deploys four distinct LLM agents, with similar task breakdowns: a planner, researcher, reporter, and judge. As of the 2025 version of EAGLE-AI core, the scoring planner and judge use o3-mini^36^, while the scoring researcher and reporter used GPT-4o^37^.

**Fig. 4.**
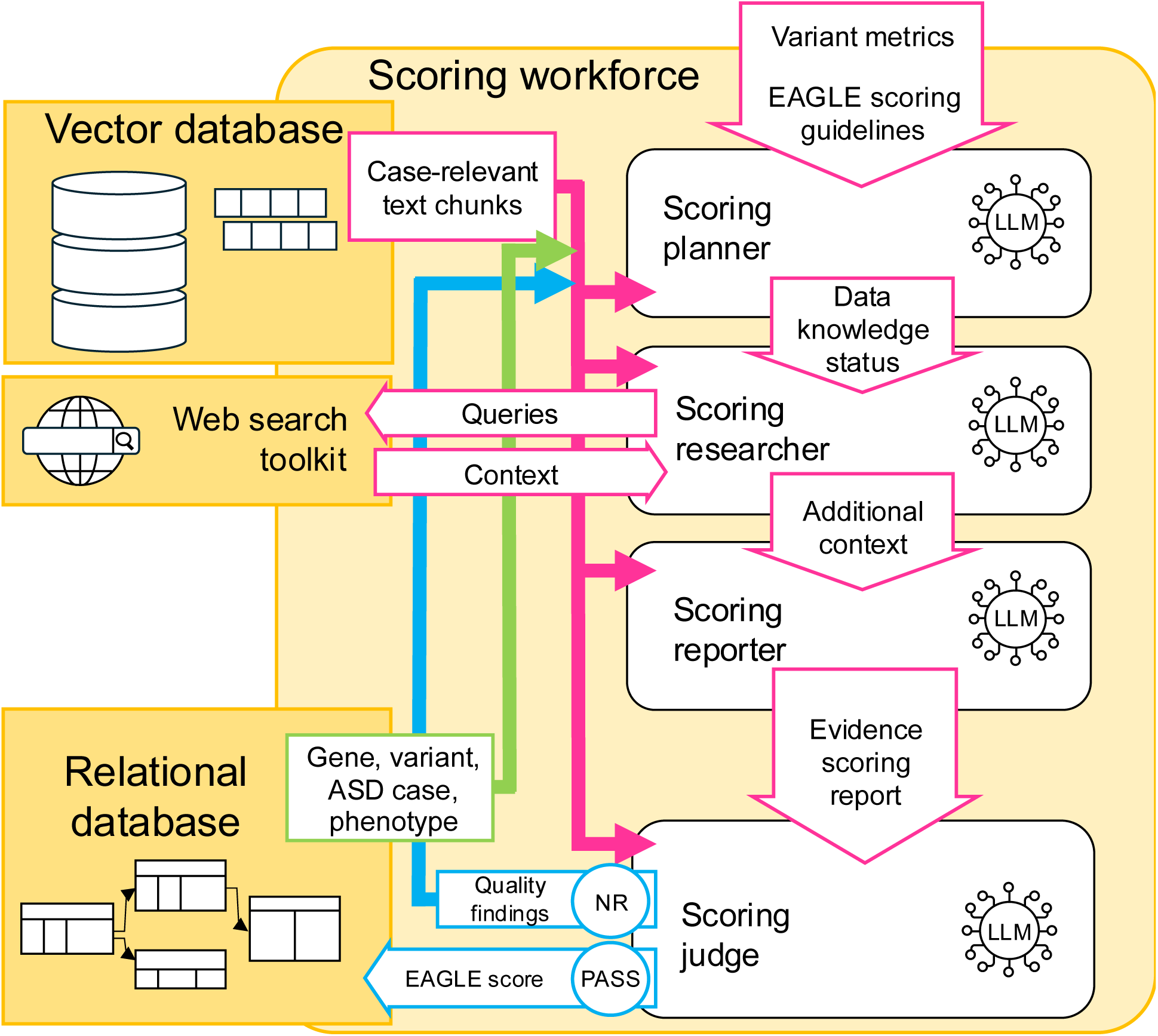
Agentic EAGLE-AI core scoring workflow. Schematic of the scoring workforce within agentic EAGLE-AI core. Initial inputs consist of case-relevant paper text chunks from the vector database; extracted gene, variant, ASD case, and phenotype data from the relational database; variant metrics from external API calls; and a prompt containing the EAGLE scoring guidelines. The workforce includes an additional task planner and coordinator agent for incorporating recommendations from the judge in the event of a “NEEDS_REVISION” (NR) verdict.

Since the EAGLE scoring guidelines (Table 1) require variant information such as null or missense status, functional data, de-novo status, and splice-site-modifying status, an upstream variant annotator agent searches for these metrics using third-party APIs (Fig. 1). The annotator sends requests to the Ensemble variant effect predictor’s (VEP)^48^ API for transcript-level, protein-level, and clinical impact; VariCarta’s^49^ for known ASD association and pathogenicity scores (CADD 1.3^50^, SIFT^51^, and polyphen^52^); gnomAD’s^53^ for population frequency; and LitVar2’s^54^ for HGVS protein variant notation and pathogenicity status. If the paper does not provide the variant protein impact, these are used in determining the impact (e.g. “missense”) for scoring purposes.

The planner agent receives the query case ID, the case’s candidate ASD-driving variant annotated with the third-party metrics, all paper text chunks relevant to the case as stored in the vector database, and all associated data from the data extraction module as stored in the relational database (Fig. 1). It receives also the EAGLE scoring guidelines (Table 1) and a checklist of data required for applying the guidelines (Table S10). Any missing data are tagged by the planner agent as requiring additional search. For all data labelled as missing, the researcher agent uses the suggested search terms and the CAMEL-AI web search toolkit to attempt to retrieve the data using a suite of search engines^46^.

A downstream reporter agent leverages all input data from the relational database, vector database, annotator, and scoring researcher in applying the EAGLE scoring guidelines as per Table 1. It returns a report for the case, summarizing the data and assigning an EAGLE score to the case’s genetic evidence, ASD phenotype evaluation, and experimental evidence (Table S11). It also summarises any evidence against the case’s ASD diagnosis or against the variant’s driving of ASD likelihood. It yields a final EAGLE score for the case, with a maximum of 2 points for genetic evidence and a maximum of 6 points for experimental evidence.

The judge agent appraises the quality of the report and issues a final verdict of “PASS” or “NEEDS_REVISION” (Table S12). In the event of a “PASS” verdict, the workforce writes the case’s EAGLE score to the relational database. In the event of a “NEEDS_REVISION” verdict, the scoring workforce incorporates the judge’s feedback and assigns follow-up tasks to the agents as appropriate (Fig. 4).

### EAGLE-AI scoring error

We tested performance for two configurations of the scoring module: the agentic scoring workforce and a deterministic if-else algorithm written in Python. The deterministic algorithm was coupled with an upstream minimal data extraction module, consisting of a BiLSTM NER model for relevance screening and a single prompt (Table S13) to GPT-4o for data extraction (see Methods).

Across 165 cases, the agentic scoring workforce achieved a case-wise mean absolute percentage error (MAPE) of 17.2% as per equation [1] (see Methods). As this is an error term, this indicates 82.8% correspondence with manual scores. Across 128 cases, the deterministic scoring algorithm achieved a case-wise MAPE of 10.8%, thereby outperforming the agentic scoring workforce by 6.4 percentage points [95% CI: −4.47, 18.47].

Distributions of scores for both the agentic workforce and the scoring algorithm closely matched that of the manually curated scores (Fig. 5a, c). For both the workforce and the algorithm, Bland-Altman plots do not indicate error bias with respect to score magnitude (Fig. 5b, d). For the agentic workforce, 85.5% of its case scores had zero difference from their corresponding manual case scores; for the algorithm, 87.5% of its case scores had zero difference.

**Fig. 5.**
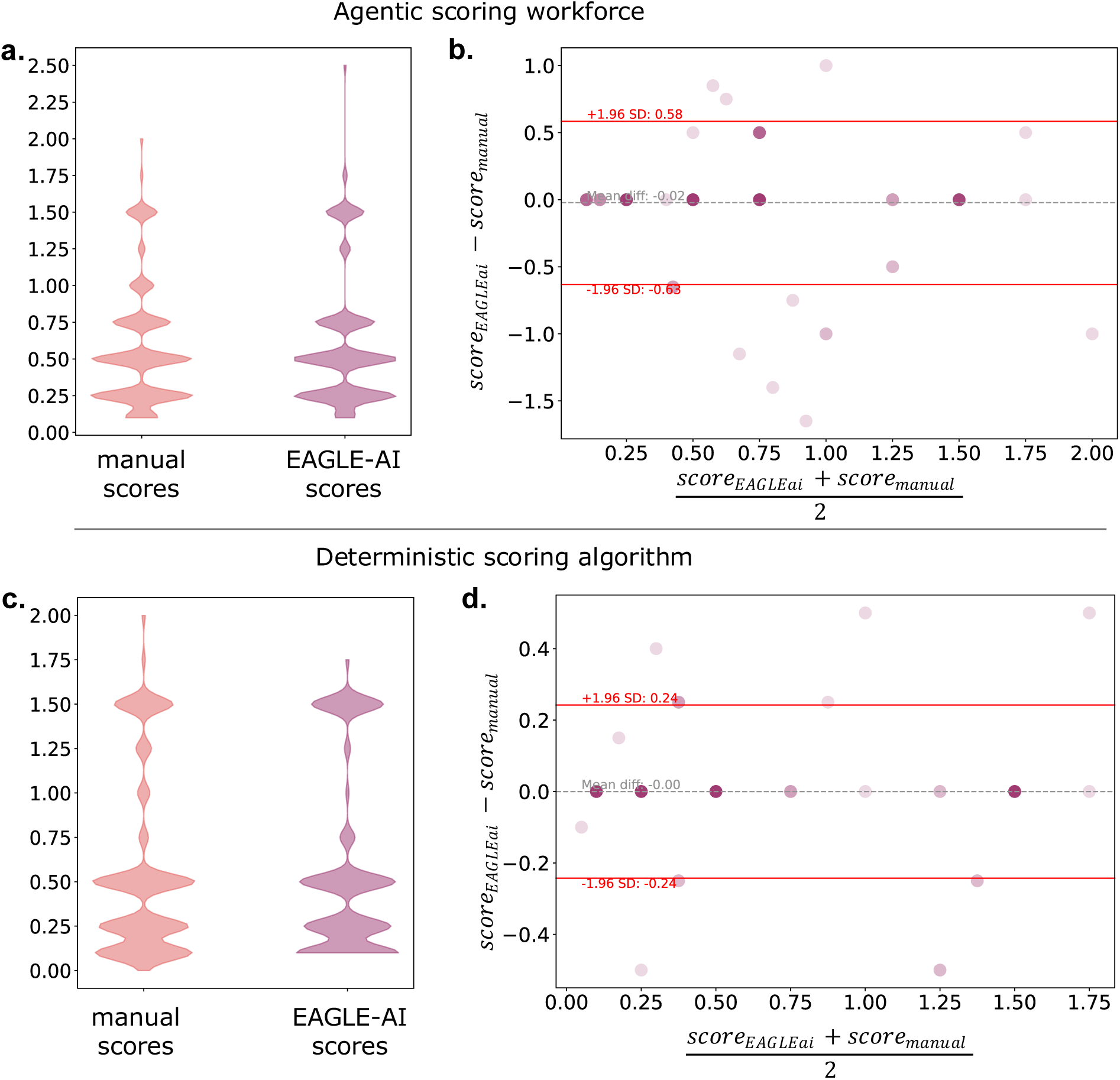
EAGLE-AI scoring performance. EAGLE-AI’s scoring performance under different conditions, where greater score means greater strength of evidence linking a case’s variant to ASD. **a, b** Agentic scoring workforce performance across 78 papers and 165 ASD cases. **a** Violin plots of manually curated case scores (orange) v. agentic (purple) EAGLE-AI case scores. **b** Bland-Altman plot for agentic EAGLE-AI’s scores benchmarked against manual scores, where darker points indicate overlap; here, each point represents an ASD case. **c, d** Deterministic scoring performance across 90 papers and 128 ASD cases. **c** Violin plots of manually curated case scores (orange) v. deterministic algorithm’s (purple) scores. **d** Bland-Altman plot for deterministic algorithm’s scores benchmarked against manual scores.

### Whole-system performance on eight genes without prior screening

We computed performance metrics for the whole EAGLE-AI system, as given by Fig. 1, using papers pertaining to eight genes that were excluded from all prior training, test, and validation sets. We used three-shot prompts to the extraction and scoring modules (Table S14, Table S15). Additionally, these papers were not screened for ease of machine readability (see Methods).The eight genes evaluated were CACNA1A, CASK, CDKL5, CHAMP1, CHD3, FMR1, GIGYF2, and GRIN1. The 36 papers contained a total of 86 candidate ASD cases with variants in these genes. Table S16 contains the failure mode annotation descriptions. Table S17 contains the human curators’ data extractions, EAGLE-AI’s data extractions, and the raw performance data. For the below, square brackets give the 95% confidence intervals.

This configuration achieved a case F1 of 71% [57%, 81%], a variant F1 of 48% [33%, 62%], and an average phenotype accuracy rating of 66 (Fig. 6a). Its scoring performance was poor, with gene-wise MAPE of 74%. We counted 10 cases whereby EAGLE-AI described cases’ co-occurring conditions in greater detail than did the manual curators; conversely, we counted 13 cases whereby the curators described them in greater detail than did EAGLE-AI (Table S17).

**Fig. 6.**
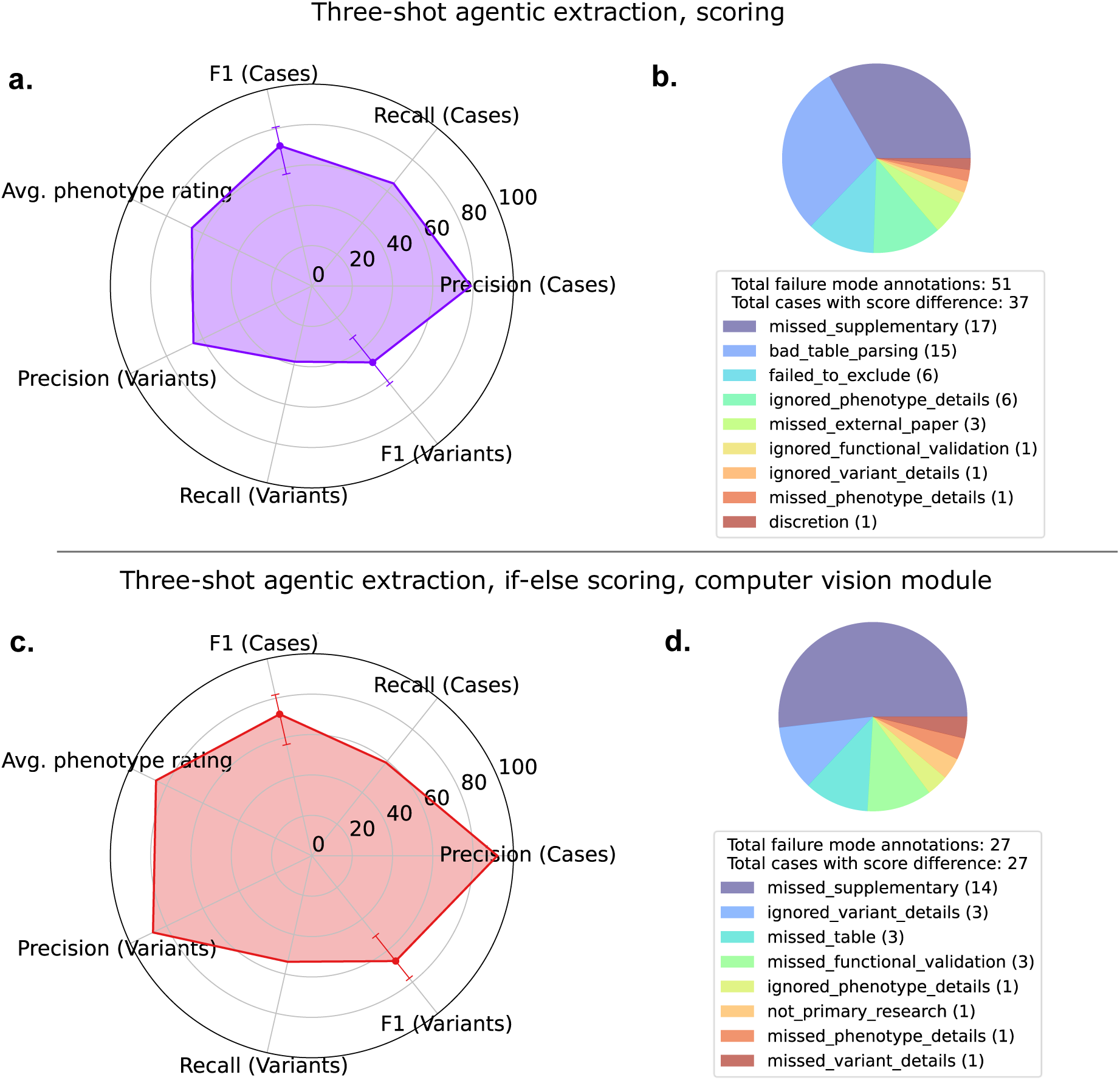
EAGLE-AI whole-system performance. EAGLE-AI’s whole-system performance metrics out of 100 using two different approaches, evaluated across manually scored cases from 36 papers about *CACNA1A*, *CASK*, *CDKL5*, *CHAMP1*, *CHD3*, *FMR1*, *GRIN1*, *GIGYF2*; error bars indicate 95% F1 confidence intervals. **a** Performance of EAGLE-AI when using three-shot-prompted agentic extraction and scoring workforces. **b** Corresponding failure-mode breakdown for all cases with scores different from that of manual curators’. **c** Performance of EAGLE-AI when using the three-shot prompted agentic extraction workforce, the if-else deterministic scoring algorithm, and a dedicated computer vision module for handling paper tables. **d** Corresponding failure mode breakdown for all cases with scores different from that of manual curators’.

To improve performance, we also pilot-tested EAGLE-AI with the if-else deterministic algorithm for scoring and a dedicated module for preprocessing and regularizing paper tables before plain-text conversion (see Methods). This configuration achieved a case F1 of 72% [56%, 82%], a variant F1 of 67% [51%, 77%], and an average phenotype accuracy rating of 86 (Fig. 6c). It achieved greater scoring performance, with gene-wise MAPE of 27%. This constitutes a scoring performance improvement of 47 [-3, 163] percentage points.

For the pilot test, the cost of running the automated curation of the 36 papers was $24.66 CAD, four labour hours, and 78 minutes of runtime. In contrast, the cost of manually curating the 36 papers was 78 labour hours.

## Discussion

On a set of 116 papers pertaining to six ASD-associated genes, EAGLE-AI achieved near-human performance on data extraction (Fig. 3). It also achieved near-human performance on scoring across 165 papers (Fig. 4). The three-shot agentic workforce, incorporating GPT-4o and o3-mini, performed best. However, the three-shot agentic, zero-shot agentic, and finetuned Llama-3.1-8B extraction modules all achieved overlapping F1 95% confidence intervals, and only the BiLSTM-CRF module significantly underperformed. Our findings serve as proof-of-concept for applying third-party LLMs, organized in multi-agent workforces, to clinical genomics evidence curation. We expect this approach will save significant amounts of time for humans.

We interpret these results with an important caveat: namely, the test papers were screened for favourable text extraction conditions. Converting the entirety of a paper to plain text is nontrivial, so we first screened these papers for ease of machine-readability (see Methods). We conducted this assessment in order to isolate performance of the data extraction and scoring modules. However, this means the input dataset did not represent the full diversity of relevant scientific literature. In particular, this excluded long papers, papers with landscape-oriented tables, papers that report critical case and variant data solely in tables, and papers with many supplementary materials. As these types of papers feature prevalently in the high-impact clinical genomics corpus, this is a limitation of the initial assessment round.

During this initial round of performance assessment, we treated human curation as the gold standard. For this reason, instances of super-human performance, whereby EAGLE-AI avoids errors made by human curators, were not possible to detect. The reported performance metrics of F1 93% and scoring MAPE 17.2% may therefore understate EAGLE-AI’s true performance on these tests.

On follow-up, whereby we evaluated the whole-system performance on eight genes without prior screening of the papers, the assessment was manual (see Methods). Here, we observed 10 cases whereby, on review, EAGLE-AI extracted correct details about ASD case phenotypes that the manual curators missed. In all such instances, the details pertained to co-occurring conditions and test results (e.g. anatomical abnormalities, brain imaging findings) (Table S17).

Under the rating system used for assessing phenotype extraction performance, these contributed only 10 out of 100 points to the phenotype accuracy rating (see Methods). We therefore updated the manually curated descriptions with EAGLE-AI’s findings and continued to treat the curations as the gold standard. Moreover, the human curators nonetheless extracted more phenotype details that were missed by EAGLE-AI than vice versa (Table S17). This finding suggests that, for extracting long lists of phenotype details and clinical test results, the combined output of EAGLE-AI and manual curators captures more correct information than either on their own. We did not observe any additional instances of super-human performance from EAGLE-AI.

The whole-system evaluation without paper screening exposed EAGLE-AI’s limitations. On this test, agentic data extraction performed worse, with final gene-wise scoring error of 74%. A follow-up failure mode breakdown indicates that errors in table parsing and errors in scoring logic contributed substantially to the reduced performance (Fig. 6b). In particular, many papers present their cases and ASD-driving variants in large tables within the full-text. Since these tables are coded as images within PDFs, EAGLE-AI relies on GPT-4o image-to-text conversion to read the table contents, which is error-prone. As per the failure mode analysis, this contributed to 15 of 37 (41%) inaccurately scored cases.

Errors in scoring logic (Table S16: failed_to_exclude, ignored_phenotype_details, ignored_functional_ validation, ignored_variant_details) contributed to 14 of 37 (38%) inaccurately scored cases. Here, agent context overload likely contributed. As the 36 follow-up papers were not screened, EAGLE-AI more often encountered critical missing information and depended on background knowledge and the web-search toolkit more. The quality of information from web search was variable and had the potential to confuse the scoring workforce. For instance, when scoring *FMR1 —* a gene canonically associated with Fragile X Syndrome, which is accompanied by autism in about 40% of male and 15% of female carriers^56^ *—* EAGLE-AI’s treated Fragile X Syndrome diagnoses as equivalent to ASD. This in turn yielded erroneous scores for many of the *FMR1* cases.

On a test set of 90 papers, our deterministic scoring algorithm performed comparably to the agentic scoring workforce and achieved a lower scoring error of 10.8%. EAGLE’s scoring framework is rigorously rules-based^10^ such that we could viably program it with if-else algorithmic logic. While LLMs are well suited for extracting data of a standardized format from scientific literature, this finding supports using traditional rules-based algorithms for evidence scoring instead of LLM agents, as if-else algorithms are not prone to the problem of context overload, hallucination, drift, or token limits.

To address the table parsing and scoring logic errors that surfaced during the whole-system evaluation on unscreened papers, we pilot-tested a new version of EAGLE-AI that uses a dedicated computer vision module for table handling and the deterministic if-else algorithm for evidence scoring (see Methods). This version of the tool achieved considerably better metrics, with average phenotype accuracy rating of 86 and gene-wise scoring error of 27%. The accompanying failure mode breakdown indicates no instances of bad table parsing and fewer instances of scoring logic errors (Fig. 6d), suggesting that the computer vision module and if-else scoring algorithm are exerting their intended effects. In light of this finding, we plan to deprecate the agentic scoring workforce on all future versions of EAGLE-AI.

Both versions of EAGLE-AI that we whole-system tested on unscreened papers showed low case recall and low variant recall (Fig. 6a, c). As indicated in the failure mode breakdowns, this was largely due to EAGLE-AI missing access to supplementary tables (Fig. 6b, d), which were sometimes the sole source of case or variant information in a paper. Papers-Nexus lacks a dedicated logic for retrieving supplementary materials, which is an area for future development. Moreover, supplementary materials sometimes contain hundreds or thousands of variants, which overwhelms the data extraction workforce by overrunning token limits during iterative prompt submission. The EAGLE curation rules also assume that authors are publishing only those variants that they view as ASD-driving: when authors present raw and unfiltered case-variant information in large supplementary tables, this breaks that assumption and leads to inflated evidence scores. Programmatic access to and handling of supplementary materials is EAGLE-AI’s principal unsolved technical problem, and we intend to address it in future releases.

Together, these limitations indicate that, even with theoretically perfect prompt engineering and model intelligence, LLMs do not entirely solve the problem of automated literature curation. Rather, LLM-mediated data extraction must be accompanied by dedicated modules for retrieval of papers, for retrieval of supplementary materials, and for PDF-to-plain-text conversion.

This points to a more inherent limitation: due to restrictions to machine access, EAGLE-AI cannot comprehensively cover the English-language literature without human intervention. The Papers-Nexus retrieval rate of 66.3% is principally limited by paywalled publishing and a lack of streamlined pathways for automated retrieval. Most journals do not support programmatic access. Currently, human curators may workaround this by manually retrieving papers and uploading them directly to EAGLE-AI via a frontend interface. In our current work, we use EAGLE-AI to regularly assess genes for their role in ASD; this includes genes emerging from our own discovery research and those added to the SFARI database^29^. Therefore, the database must be updated regularly, and human retrieval and uploading incurs greater ongoing labour costs associated with running the workflow. As of now, this semi-automated approach with EAGLE-AI remains untested at a large scale.

EAGLE-AI requires ongoing maintenance to ensure stable performance. We update our API calls as new LLMs are released: currently, EAGLE-AI uses GPT-5. We monitor LLM performance drift by evaluating gene-wise scoring SMAPE on a small set of test papers.

Setting aside the current performance limitations, we propose that EAGLE-AI’s architecture can generalize to clinical genomics evidence curation as a whole. For instance, EAGLE-AI’s workflow may be able to replace or augment the manual curation approach used by ClinGen’s gene-disease validity working groups. During the whole-system pilot test of if-else scoring and the computer vision module, our cost measurements indicate that EAGLE-AI can save an average of 1.5 to 2 human labour hours per paper.

Some features of EAGLE-AI are tailored to the EAGLE curation task. However, we designed EAGLE-AI with modularity in mind. To apply the workflow to a different curation task, for example to curating for a different disease or disorder, much of the architecture is reusable. For a workflow that performs only data extraction, one would need to change only the search terms, prompts to the data extraction workforce, and schema of the relational database. For each new curation task, however, the prompts and schema would require design consultation with both manual curators and domain subject-matter experts.

For additional evidence scoring based on ClinGen’s semi-quantitative (“Definitive”, “Strong”, “Moderate”, “Limited”) framework^13^, a tailored scoring algorithm would likely need to be developed for each genetic disease or condition. For curation tasks whereby scoring requires considerable subjective decision-making, an agentic LLM workforce may prove more suitable. However, one might re-apply a suite of general principles (e.g. nonsense de-novo variants and deletions present stronger evidence for a monogenic role than inherited or missense variants), such that the development time for disease-specific scoring modules may be short.

The generalizability of EAGLE-AI and other LLM-based curation tools bears implications regarding intellectual property best practices. To conduct data extraction, workflows like EAGLE-AI must send paper contents to external APIs, whereby third parties may retain the paper contents and subsequently use them for training their own models. Here, patient privacy is not as much a concern because best practices in publishing already protect personal health information and the papers are already publicly available. We limited intake papers to those published under open access, which permits broad dissemination and reuse of scientific literature. However, open access licensing was not designed with LLM curation in mind, and authors do not necessarily consent to the transmission of full-text papers to proprietary APIs. Additionally, authors’ ownership over their paper contents must be weighed against a responsibility to transparently and accessibly disseminate the findings of publicly funded research. Given that LLM curation workflows pose challenges to intellectual property restrictions while standing to greatly facilitate dissemination and accessibility, licensing options for scientific literature must evolve to account for the emerging prevalence of LLMs.

With the advent of LLMs, we expect that automated workflows for evidence curation will become more common. As a result, the human labour of curating evidence may transition away from manual data extraction or evidence scoring and toward designing database schemas, LLM prompts, and evidence scoring algorithms. Moreover, researchers and publishers may come to reconsider how scientific material is published, for example by more often supporting open-access and machine-readable formats.

## Conclusions

Here we present proof-of-concept for EAGLE-AI, which automates the majority of a clinical genomics literature curation workflow for ASD. This includes the data extraction, evidence scoring, and database entry steps. The tool achieves near-human performance on screened literature. However, whole-system testing on unscreened papers reveals limitations due to LLM context overload, table parsing, and handling of supplementary materials. Though programmatic retrieval and handling of supplementary data remains an unsolved limitation, pilot testing shows that an if-else scoring algorithm and dedicated computer vision module resolves the scoring context overload and table parsing problems.

## Methods

### Named entity recognition (NER) model architecture

With the aim of benchmarking the LLM workforce against traditional machine learning methods, we developed a version of EAGLE-AI core that does not use LLM agentic workforces but rather depends on a bilateral long short-term memory (BiLSTM) neural network with a conditional random-field (CRF) layer. Using PyTorch^61^, we trained the BiLSTM-CRF model to perform named entity recognition (NER) of genes, variants, and medical condition terms from the extracted literature text. The entity mentions are used to relevance-filter and assemble text chunks before loading them into the EAGLE-AI data extraction module. Here, the data extraction module queries a single GPT-4o API endpoint^37^ for data extraction or, alternatively, a finetuned Llama 3.1-8B model^47^.

The NER model consists of an embedding layer, an attention layer, a BiLSTM layer, and a CRF layer. We used pretrained BioWordVec embeddings^62^ for the embedding layer, with a vocabulary size of 10,475 and where each embedding vector is 200-long. Each embedding vector is concatenated to a corresponding 150-long output from a character encoder, consisting sequentially of a linear layer and ReLU activation function. Both the embedding layer and the character encoder were further trained alongside the rest of the model.

The BiLSTM layer consists of two feed-forward LSTM networks with 192-long hidden states. Its output is a tensor of emission scores, which feed into an attention layer consisting sequentially of a linear layer, Tanh activation, multiplication by a context vector, and SoftMax activation. Output from the attention layer feeds into the CRF layer; the CRF layer also receives a target tensor, which contains the true classification scores as per the labelled train/test data. The NER model labels each token as one of 31 named entities pertaining to case, variant, and phenotype information.

### NER model training

We trained the NER model in PyTorch^61^ using the BioRED dataset, which consists of 600 clinical PubMed abstracts^63^. Tokens were generated by first splitting the text into sentences by punctuation boundaries, then separating words by whitespace and isolating leading or trailing punctuation marks as individual tokens. Here, the labelled entities of interest pertained to genes, genomic variants, diseases/disorders, patient case information, signs/symptoms, and experimental validation. We converted the labelling scheme from beginning, inside, outside (BIO) to a binary entity labelling scheme whereby each token is labelled as either inside an entity or outside any entity. We treated output of the CRF layer as the loss function. The model was trained for 36 epochs at a learning rate of 2e-2 and with dropout of 0.5.

### Llama 3.1-8B finetuning

With the aim of benchmarking an LLM workforce against a single finetuned LLM, we conducted chat-formatted supervised finetuning of a single Llama 3.1-8B model^47^.

The finetuning set consists of 23,627 tasks drawn from 297 distinct papers, with 254 papers constituting the training set and 43 papers constituting the evaluation set (85:15 split). Each task consists of a query, with instructions and a plain-text paper excerpt, and a target response. For full case extraction tasks, the target response is a case JSON with sex, age, phenotype, and variant fields. The dataset includes smaller-scale target responses for identifying a case’s intellectual ability, phenotype confidence, variant type, and variant de novo status.

Text was tokenized using LlamaTokenizerFast. Using a low-rank (LoRA) matrix finetuning approach, we targeted parameters in the query, key, value, and output projection tensors of the model’s attention mechanism, as well as the gate, up, and down projection tensors of the multilayer perceptron block. Finetuning was conducted over three epochs using the SFTTrainer from the HuggingFace transformer reinforcement learning library^64^. We used a cross-entropy loss function applied exclusively to the assistant response tokens and a learning rate of 2e-4. For parameter optimization, we used paged AdamW 32-bit.

### Data extraction precision-recall evaluation

In testing performance of EAGLE-AI core’s data extraction precision-recall, we drew from evidence extracted by EAGLE’s manual curators. Here, the aim was to test the data extraction step in isolation. We constrained the cases to only those that had a final EAGLE score and had a reported variant in *DMD, SHANK1, CACNA1D, DDX3X, DDX53*, or *MECP2*.

We additionally screened papers for ease of machine-readability. Here, papers were excluded if they exceeded 128,000 tokens after processing with the o200k_base tokenizer^37,38^, if they included landscape-oriented tables, if they reported critical case or variant information solely in tables, if they included more than one supplementary table or figure, or if they included supplementary materials in formats other than markdown, plain-text, or PDF.

EAGLE’s manual curation workflow is already established and described elsewhere^10^. We collected and stored the source literature from which the manual curators extracted patient/subject information, phenotype descriptions, and variant information. We stored embeddings for the documents in EAGLE-AI’s vector database and used them as input to EAGLE-AI core.

In evaluating EAGLE-AI’s data extraction performance, we treated presence of an extraction as positive and absence of an extraction as negative. We evaluated the case ID extractions, variant ID extractions, and phenotype descriptions as separate categories. For each data extraction category, we computed the true positive, false positive, and false negative rate using the definitions in Table 2. This gave us the corresponding precision, recall, and F1-score of EAGLE-AI core’s data extractions. We determined confidence intervals for F1 scores by solving for the real roots of the Wilson indirect quartic function^65^.

We matched cases extracted by EAGLE-AI to their corresponding manually curated cases by first matching by normalized publication name and author. We achieved further case-specific matching by adding confidence scores in the event of a matching gene name, case ID, or inheritance pattern.

We tested EAGLE-AI’s multi-agent workforce when using task prompts with zero input-output examples (“zero-shot”) (Fig. 3c) and three input-output examples (“three-shot”) (Fig. 3d, Table S14), respectively. We also tested data extraction precision-recall of the NER model when used for relevance filtering and text chunk assembly for a given case: during testing, we coupled the NER model with either a single GPT-4o API endpoint^37^ for data extraction (Fig. 3a) or the finetuned Llama 3.1-8B model for data extraction (Fig. 3b).

### If-else deterministic scoring algorithm

As an alternative to the agentic scoring workforce, we wrote a deterministic scoring algorithm that applies if-else logic in Python. The algorithm applies the rules-based EAGLE scoring guidelines for genetic, phenotypic, and experimental evidence as described in Table 1. However, because if-else logic cannot program subjective decision-making, the algorithm applies only the default scores instead of the discretionary score ranges specified in brackets.

In the deterministic script, a ScoreConstants class stores the default scores given in Table 1A. Here, the extraction schema given by Table S1 was modified for easier handling by the downstream deterministic scoring algorithm. The algorithm instead reads from three distinct schemas populated by the data extraction workforce: one for genetic evidence, one for phenotypic evidence, and one for experimental evidence.

The genetic evidence schema is condensed to case_id, variant_id, rationale, and variant fields; the variant field contains subfields indicating the inherited pattern (i.e. de novo or inherited) and protein-level impact (e.g. “Missense”, “Stop gain”, “Splice site”). The phenotypic evidence schema gives a dedicated field for phenotypic confidence as “high”, “medium”, or “low”. The schema for experimental evidence stores the type of experimental evidence if present, e.g. “Rescue in non-human model organism” or “Protein Interaction assay”.

Subsequent logic assigns the protein impact term to one of three buckets: null, canonical splice, and non-canonical splice. Using the bucket assignment, a de novo scoring function (_score_inherited_variant) applies the first two rows of Table 1A to de novo variants, whereas an inherited scoring function (_score_inherited_variant) applies the third and fourth rows to inherited variants. This gives the genetic evidence score, which is passed into a phenotype scoring adjustment function (_calculate_phenotype_adjustment) alongside the phenotype confidence level, thereby programming the logic in the bottom half of Table 1B. For cases with missing critical data for genetic evidence or phenotypic evidence, these functions assign a final score of 0 to the case. A function for handling the functional evidence (_assess_functional_evidence) applies the logic in Table 1C.

**Table 1A:**
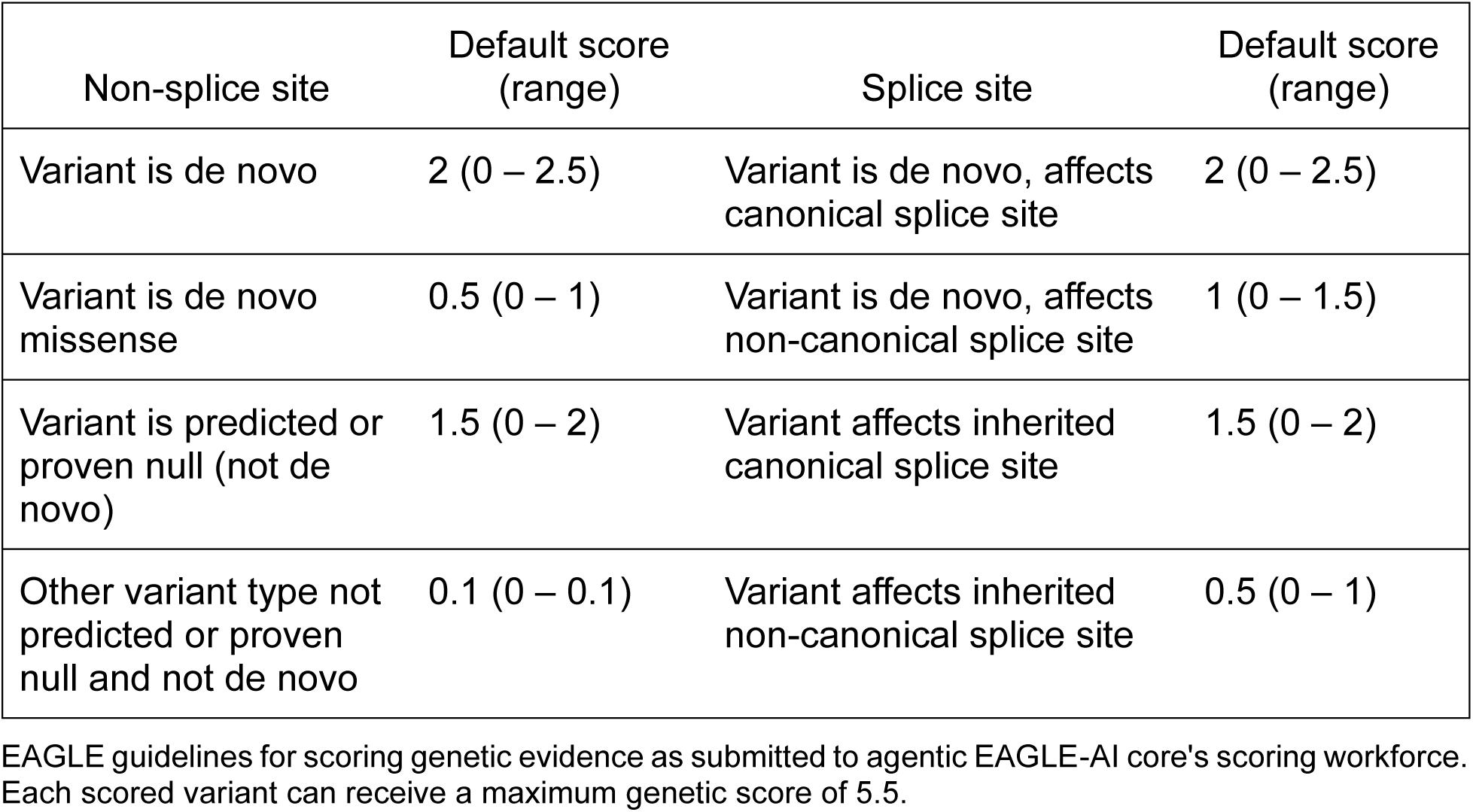
EAGLE scoring guidelines, genetic evidence.

| Non-splice site | Default score<br>(range) | Splice site | Default score<br>(range) |
| --- | --- | --- | --- |
| Variant is de novo | 2 (0 – 2.5) | Variant is de novo, affects canonical splice site | 2 (0 – 2.5) |
| Variant is de novo missense | 0.5 (0 – 1) | Variant is de novo, affects non-canonical splice site | 1 (0 – 1.5) |
| Variant is predicted or proven null (not de novo) | 1.5 (0 – 2) | Variant affects inherited canonical splice site | 1.5 (0 – 2) |
| Other variant type not predicted or proven null and not de novo | 0.1 (0 – 0.1) | Variant affects inherited non-canonical splice site | 0.5 (0 – 1) |

EAGLE guidelines for scoring genetic evidence as submitted to agentic EAGLE-AI core's scoring workforce. Each scored variant can receive a maximum genetic score of 5.5.

**Table 1B:** EAGLE scoring guidelines, ASD phenotype evaluation.

| Confidence level | Indicators |
| --- | --- |
| High confidence | Expert clinician diagnosis, validated assessment methods, or explicit DSM or ICD criteria mentioned |
| Medium confidence | Description of symptoms in social / communicative and repetitive domains indicative of ASD |
| Low confidence | Mentions only "ASD", "autism", "autistic features", "autistic traits", etc. |

| If low confidence, apply deduction ... |  | If medium confidence, apply deduction ... |  |
| --- | --- | --- | --- |
| genetic evidence score | deduction | genetic evidence score | deduction |
| 2 | -1 | 2 | -0.5 |
| 1.5 | -0.5 | 1.5 | -0.25 |
| 1 | -0.5 | 1 | -0.25 |
| 0.5 | -0.25 | 0.5 | -0.1 |

EAGLE guidelines for scoring ASD phenotype evaluation as submitted to agentic EAGLE-AI core's scoring workforce. Each case receives a confidence level of high, medium, or low. If the confidence level is medium or low, a downstream algorithm performs a deduction to the genetic evidence score, depending on the initial genetic evidence score. If the case cannot meet the minimum confidence level (low), the case is excluded.

**Table 1C:** EAGLE scoring guidelines, experimental evidence.

| Evidence category | Evidence type | Suggested points default | Suggested points range | Max score |
| --- | --- | --- | --- | --- |
| Function | Biochemical function | 0.5 | 0 – 2 | 2 |
|  | Protein interaction | 0.5 | 0 – 2 |  |
|  | Expression | 0.5 | 0 – 2 |  |
| Functional alteration | Patient cells | 1 | 0 – 1 | 4 |
|  | Non-patient cells | 0.5 | 0 – 4 |  |
| Models | Non-human model organism | 2 | 0 – 4 | 4 |
|  | Cell culture model | 1 | 0 – 2 |  |
| Rescue | Rescue in human | 2 | 0 – 4 | 4 |
|  | Rescue in non-human model organism | 2 | 0 – 4 |  |
|  | Rescue in cell culture model | 1 | 0 – 2 |  |
|  | Rescue in patient cells | 1 | 0 – 2 |  |
| Total allowable points for experimental evidence |  |  |  | 6 |

EAGLE guidelines for scoring experimental evidence as submitted to agentic EAGLE-AI core's scoring workforce.

**Table 2:** EAGLE-AI core data extraction performance testing definitions.

|  |  |  |
| --- | --- | --- |
| Case ID and Variant ID extractions | Exact match after normalization | True Positive |
|  | Less than exact match | False Positive |
|  | Absent variant ID | False Negative |
| Phenotype description extractions | Embedding cosine similarity $\geq 0.7$ | True Positive |
| | Embedding cosine similarity $< 0.7$ | False Positive |
|  | Absent description | False Negative |

Definitions of true positive, false positive, and false negative used in computing precision, recall, and F1-score for case ID, variant ID, and phenotype description data extractions. We matched EAGLE-AI's data extractions to their corresponding manual ground truth using the patient/subject case ID. Variant IDs and case IDs were normalized for whitespace and punctuation. Phenotype descriptions extracted by both EAGLE-AI and the manual curators were converted to embeddings using the OpenAI text-embedding-3-large matrix. Matches were assessed in reference to ground truths produced by manual EAGLE curators.

### Evidence scoring error evaluation

We tested performance of the agentic scoring workforce when provided with three correct scoring examples in the initial task prompt (“three-shot”) (Table S15).

Additionally, we measured performance of the deterministic if-else scoring algorithm written in Python. We coupled the algorithm with the BiLSTM-CRF NER model, which was used for relevance-screening and assembling text chunks. To generate the data extractions, we used post-NER regular expression matching to augment entity labelling for the purposes of relevance filtering. We then submitted a single dynamic prompt to a GPT-4o-mini endpoint^38^, which instructed the model to extract the data (Table S13). We used the resulting extractions as input to the deterministic algorithm.

In evaluating EAGLE-AI core’s scoring error, we computed the mean absolute percentage error (MAPE) as per equation [1]. The aim was to evaluate performance of the scoring step in isolation. Here, *n* denotes the number of genes when computing gene-wise MAPE or the number of candidate ASD cases when computing case-wise MAPE.

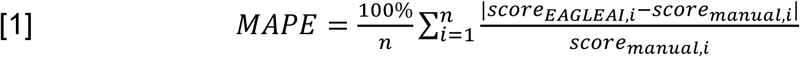

We matched cases scored by EAGLE-AI to their corresponding manually curated cases by first matching by normalized publication name and author. We achieved further case-specific matching by adding confidence scores in the event of a matching gene name, case ID, or inheritance pattern. For each case extracted by EAGLE-AI, we treated its highest-confidence match as the corresponding manually curated case when applying equation [1].

We evaluated the scoring error of EAGLE-AI’s multi-agent workflow when given zero-shot and three-shot task prompts.

We also evaluated scoring performance of EAGLE-AI when using the NER model augmented by regular expression pattern matching and coupled to a single GPT-4o-mini endpoint^38^ for data extraction using the prompt in Table S13. Here, scoring was performed using the deterministic if-else algorithm as per the rules in Table 1.

### Whole-system evaluation on eight genes without prior screening

We evaluated performance of the whole EAGLE-AI system using the three-shot agentic LLM workforces for the data extraction and scoring steps (Fig. 6a, b). During three-shot scoring, the prompt to the scoring reporter agent was scaled down to avoid context overload (Table S15). Instead of a five-section markdown report, the reporter agent was instructed to provide only a four-line rationale accompanied by the evidence score fields.

The aim was to evaluate performance of the entire workflow, including retrieval of supplementary materials, paper screening, data extraction, scoring, and database incorporation. The genes evaluated were *CACNA1A*, *CASK*, *CDKL5*, *CHAMP1*, *CHD3*, *FMR1*, *GIGYF2*, and *GRIN1*. These genes were selected because they were never before used in designing, training, or testing EAGLE-AI and were never scored using manual EAGLE curation. Moreover, some have strong evidence for association with ASD as per the SFARI database^29^, while others have relatively little evidence in the literature. We included 36 papers related to these genes. Papers were included only if they were successfully downloaded by Papers-Nexus and assigned to these genes by the gene flagger module. The papers were not screened for ease of machine-readability.

We evaluated EAGLE-AI’s performance across 86 candidate ASD cases extracted by manual curators from the papers. The manual curators extracted the gene name, paper name, case ID, phenotype description, phenotype confidence level, sex of the case, age of the case, variant ID, zygosity of the variant, variant inheritance (i.e. de-novo or inherited), and variant impact (e.g. missense). The manual curators also assigned evidence scores to each of the cases using the EAGLE standard operating procedures (Table 1). To control against variability due to subjective decision making, the curators applied only the default scores rather than the suggested score ranges. The curators received the 36 papers and 8 gene names but were blinded to EAGLE-AI’s data extractions and scores for the 86 cases. Data extraction and scoring were not performed in duplicate.

We measured EAGLE-AI’s performance by manually matching EAGLE-AI’s output to the output of the curators. We matched cases by case ID; in the absence of a matching case ID, we matched cases by phenotype description, age of the case, sex of the case, and/or paper name. We matched variants by rsID^66^, ClinVar ID^67^, HGVS NM number and coding position, HGVS NC or NG number and genome position, HGVS NP number and protein position^68^, or chromosome:position:ref>alt notation relative to a reference genome. We designated matches as true positive (TP), false positive (FP), false negative (FN), or true negative (TN).

Sometimes, a case and variant extracted by the manual curators matched to a case extracted by EAGLE-AI, but EAGLE-AI reported an incorrect variant. Under these conditions, we counted the variant as both FP and FN. From the count of TPs, FPs, FNs, and TNs, we computed the precision, recall, and F1 score out of 100 percent for both the cases and variants. For F1, we determined 95% CIs by solving for the real roots of the Wilson indirect quartic function^65^.

For each of the 86 cases, we measured the phenotype accuracy using a tailored 100-point rating system. If EAGLE-AI was correct about ASD status, it received +50 points. If EAGLE-AI was correct about ASD confidence level and reason, it received +20 points. If EAGLE-AI was correct about major co-occurring conditions, i.e. epilepsy, seizures, intellectual disability, and developmental delay, it received +20 points. If EAGLE-AI was correct about detailed co-occurring conditions and testing results, e.g. other psychiatric disorders, electroencephalogram results, brain imaging results, micro or macrocephaly, or anatomical abnormalities, it received +10 points. The rating system was designed a priori by one of the manual curators and reflects their sense of each feature’s relative importance when extracting case phenotypes.

For each of the eight genes, we also measured the gene-wise scoring error as per equation [1], where both *score_EAGLEAI-i_* and *score_manual,i_* are summed across all cases for a given gene.

For all cases whereby EAGLE-AI assigned inaccurate final scores (*n* = 37), we annotated the reason(s) for inaccuracy (i.e. the failure mode(s)), which are described in Table S16. Failure modes related to the extraction module begin with “missed_”, whereas errors related to the scoring module begin with “ignored_” or “failed_”. 14 cases had two contributing failure modes, giving 51 total annotations. We counted the number of times each failure mode was observed (Fig. 6b).

We noticed that EAGLE-AI sometimes described conditions co-occurring with ASD in greater detail than did the manual curators. Since this was a potential instance of superhuman performance, we counted the number of times this occurred, as well as the number of times the curators described co-occurring conditions in greater detail than did EAGLE-AI (Table S17).

### Whole-system pilot test with if-else scoring algorithm, computer vision module

To improve performance, we pilot tested EAGLE-AI’s whole system while using the deterministic if-else algorithm for evidence scoring (Fig. 6c, d). Our aim in using the if-else algorithm was to reduce the count of scoring logic errors, i.e. those whose annotations begin with “ignored_” or “failed_”, during whole-system evaluation. To reduce the count of bad_table_parsing errors, we used a dedicated computer vision module for pre-processing and regularizing paper tables and converting them to text. This was applied instead of table-to-markdown conversion with GPT-4o, which we found to be error-prone.

The computer vision module first handles each page of an input PDF and detects its orientation. Here, OpenCV is used to detect the relative frequency of horizontal vs. vertical lines (morphologyEx(), findContours(), HoughLinesP())^55^, whereas Tesseract is used to determine glyph orientation when possible (image_to_osd())^71^. If the page is rotated, the module reorients it. This regularizes tables for easier conversion downstream. Docling^72^ is then used to identify PDF sections containing tables; it calls to the IBM TableFormer model to build a markdown skeleton of table lines, which is then aligned to the text of individual table cells. If the cell contents are coded as images rather than text, EasyOCR^73^ performs the pixel-to-text conversion.

For this pilot test, we used the same 36 papers and 86 cases used for the previous whole-system evaluation with *CACNA1A*, *CASK*, *CDKL5*, *CHAMP1*, *CHD3*, *FMR1*, *GIGYF2*, and *GRIN1*. The protocol for measuring performance on case extraction, variant extraction, phenotype extraction, and scoring was the same. We additionally measured the monetary, labour-hours, and runtime cost of the automated curation with EAGLE-AI. The cost measurement did not include hardware, electricity, or development.

## Ethics approval and consent to participate

This study was approved by The Hospital for Sick Children Research Ethics Board (1000080561).

## Consent for publication

Not applicable: all case reports contained in the present study are from published literature.

## Availability of data and materials

Much of the data generated and analyzed during the current study are included in the published article and its supplementary information files. Complete datasets for the performance evaluations, model finetuning, and model training are available at the EAGLE-AI proof-of-concept data repository at https://github.com/juliandeanmoran/EAGLE-AI_poc_publicData. Code for the performance assessments, subsequent analyses, and deterministic if-else algorithm is also available at the public repository.

## Authors’ contributions

VF developed Extractify, the gene flagger module, the search endpoint of Papers-Nexus, and all versions of EAGLE-AI core; he conducted bulk automated performance assessments of all EAGLE-AI core data extraction and scoring module versions. JM developed the PDF-miner endpoint of Papers-Nexus, conducted manual curation and follow-up performance assessment for agentic EAGLE-AI on eight novel genes, tested and debugged EAGLE-AI core and Extractify, assisted in automated performance assessment of the scoring modules, and was the principal writer of the manuscript. NBS and OR manually curated EAGLE-AI’s ground truth datasets and tested and gave feedback on EAGLE-AI; NBS assisted in the manual performance assessment of EAGLE-AI on eight genes. VF, OR, and NH contributed to writing the manuscript. AW contributed to developing Papers-Nexus. VF, MMA, and WE initiated the project and conceptualized the framework. JASV and SWS provided supervision, funding acquisition, oversight, and guidance throughout the project. All authors read, gave feedback on, and approved the manuscript.

## Acknowledgements

We wish to acknowledge the following resources: MSSNG (www.mss.ng), by Autism Speaks and The Centre for Applied Genomics at The Hospital for Sick Children, Toronto, Canada; and SPARK (www.sparkforautism.org), by the Simons Foundation Autism Research Initiative. We also thank the participating families for their time and contributions to these databases, as well as the generosity of the donors who supported these programs. Additionally, we wish to thank all research authors whose data published under open access enabled development of EAGLE-AI.

## Supplementary figures and tables

**Table S1:** EAGLE-AI core’s data extraction schema with example values.

| Field | Example value |
| --- | --- |
| case_id | "CASE123" |
| description | "Male aged 0-5 years diagnosed with ASD at age 0-5 years, showing delayed speech development and repetitive behaviors. Genetic testing revealed a pathogenic variant in the SHANK3 gene." |
| variant_extracted | True |
| variant | "chr22:51159835G>A" |
| age | "0-5 years" |
| sex | "male" |
| ethnicity | "caucasian" |
| family_history | "No family history of ASD or related disorders" |
| phenotype_confidence | "High" |
| diagnostic_criteria_used | "DSM-5" |
| expert_clinical_diagnosis | True |
| multidisciplinary_team | True |
| validated_assessment_methods | True |
| core_asd_symptoms | "Limited social communication, repetitive behaviors, sensory sensitivities to loud sounds" |
| description_of_core_asd_symptoms | "Limited eye contact, delayed speech development, hand flapping when excited, and distress in noisy environments", |
| age_at_diagnosis | "0-5 years" |
| assessment_tools_used | "ADOS-2, ADI-R" |
| explicit_mention_of_dsm_icd | True |
| cognitive_assessment_results | "FSIQ 85, with relative strengths in visual processing" |
| comorbidities | "ADHD, mild intellectual disability" |
| phenotypes | "speech delay, repetitive behaviors, social communication deficit, hypersensitivity to sound" |
| contradictory_evidence | None |
| genotyping_method | "Trio WGS (Illumina NovaSeq 6000)" |
| gene | "SHANK3" |
| chromosome | "22" |
| reference_genome | "hg38" |
| position | "chr22:51159835" |
| region | "22q13.33" |
| transcripts | ["NM_033517.1"] |
| genomic_hgvs | "g.51159835G>A" |
| cdna_hgvs | "c.3679C>T" |
| protein_hgvs | "p.Arg1227Ter" |
| rsid | "rs1555569037" |
| variant_type | "nonsense" |
| variant_description | "Heterozygous nonsense mutation in SHANK3 resulting in a premature stop codon, predicted to cause loss of function" |
| zygosity | "heterozygous" |
| population_frequency | gnomAD: "0.00001"<br>ExAC: "Not observed" |
| inheritance_pattern | "de_novo" |
| segregation_data | "Variant not present in unaffected parents" |
| sift_score | None |
| polyphen_score | None |
| cadd_score | 35.2 |
| other_scores | "REVEL": 0.95<br>"MutationTaster": 0.99 |
| linkage_to_asd | True |
| impact | "nonsense (predicted pathogenic)" |
| supporting_quotes | "The patient demonstrates significant deficits in social communication, and interaction across multiple contexts, as well as restricted and repetitive patterns of behavior.",<br>"Trio exome sequencing revealed a de novo nonsense variant in SHANK3 (c.3679C>T, p.Arg1227Ter), classified as pathogenic according to ACMG guidelines." |
| additional_notes | "No standardized IQ scores available from early developmental period", "Minor discrepancy in reported age of walking milestone between parent report and medical records" |
| rationale | "Included due to the presence of a well documented de novo pathogenic variant in SHANK3, a known ASD-associated gene, along with comprehensive clinical description meeting DSM-5 criteria for ASD." |

Table representation of output instructions, expressed as an example of a populated schema, contained within the prompt to agentic EAGLE-AI core's data reporter agent. Instructions are formatted as a Python dictionary within the prompt string.

**Table S2:**
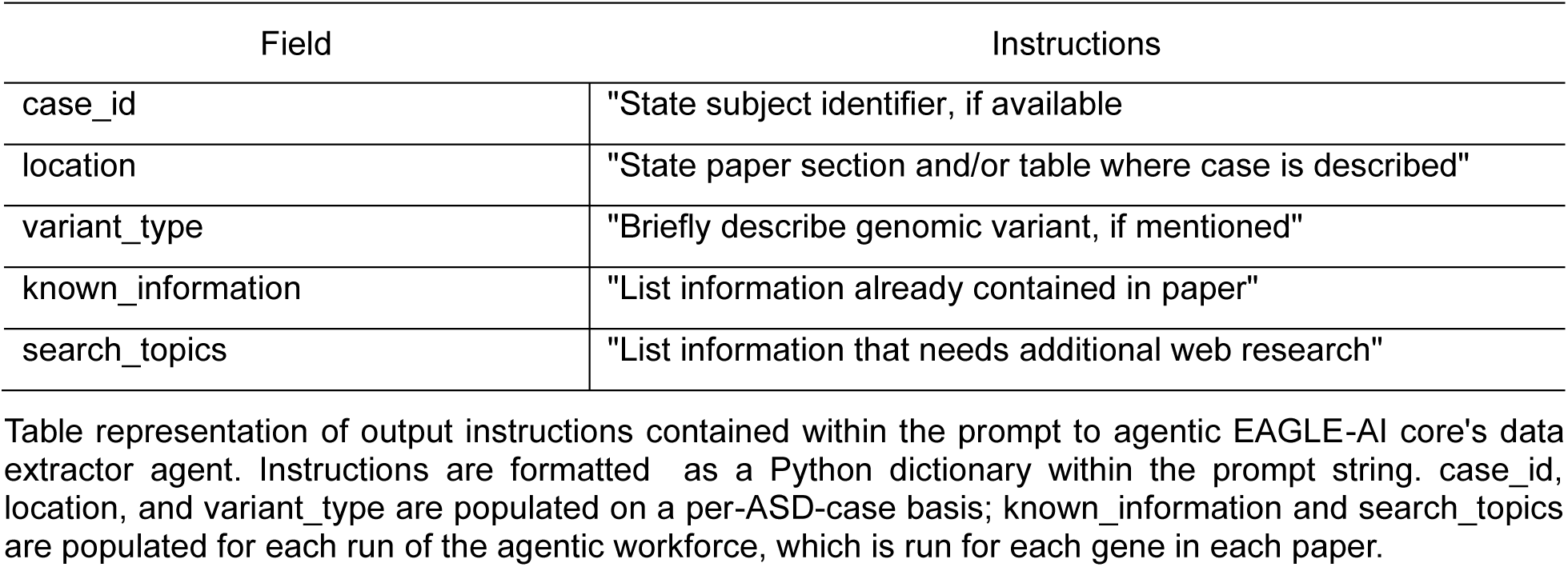
Agentic EAGLE-AI core’s data extractor output instructions.

| Field | Instructions |
| --- | --- |
| case_id | "State subject identifier, if available" |
| location | "State paper section and/or table where case is described" |
| variant_type | "Briefly describe genomic variant, if mentioned" |
| known_information | "List information already contained in paper" |
| search_topics | "List information that needs additional web research" |

Table representation of output instructions contained within the prompt to agentic EAGLE-AI core's data extractor agent. Instructions are formatted as a Python dictionary within the prompt string. case\_id, location, and variant\_type are populated on a per-ASD-case basis; known\_information and search\_topics are populated for each run of the agentic workforce, which is run for each gene in each paper.

**Table S3:** Agentic EAGLE-AI core’s data verifier output instructions.

| Field | Instructions |
| --- | --- |
| case_id | "State subject identifier, if available" |
| location | "State paper section and/or table where case is described" |
| variant_type | "Briefly describe genomic variant, if mentioned" |
| reason_missed | "State reason for why this case might have been missed" |
| extraction_priority | "Give as High, Medium, or Low" |
| additional_findings | "State additional findings from web search" |
| verified_information | "State information verified by web search" |

Table representation of output instructions contained within the prompt to agentic EAGLE-AI core's data verifier agent. Instructions are formatted as a Python dictionary within the prompt string. case\_id, location, variant\_type, extraction\_priority, and reason\_missed are populated on a per-ASD-case basis; additional\_findings and verified\_information are populated for each run of the agentic workforce, which is run for each gene in each paper.

**Table S4:** Agentic EAGLE-AI core’s data judge output instructions.

| Field | Instructions |
| --- | --- |
| evaluation_complete | "Give as True or False" |
| format_adherence | "Give as Complete, Partial, or Incomplete" |
| data_completeness | "Give as Complete, Partial, or Incomplete" |
| critical_missing_fields | "List any critical missing fields" |
| overall_quality | "Give as High, Medium, or Low" |
| consistency_issues | "For each case with consistency issue(s), give the case_id, describe the consistency issue(s), and provide a recommendation for how to resolve it" |
| final_verdict | "Give as 'PASS' or 'NEEDS_REVISION'" |
| recommendations | "List specific recommendations for improving the data extraction report" |

Table representation of output instructions contained within the prompt to agentic EAGLE-AI core's data judge agent. Instructions are formatted as a Python dictionary within the prompt string. final\_verdict and recommendations are considered by the CAMEL workforce's coordinator and task planner agents.

**Table S5:**
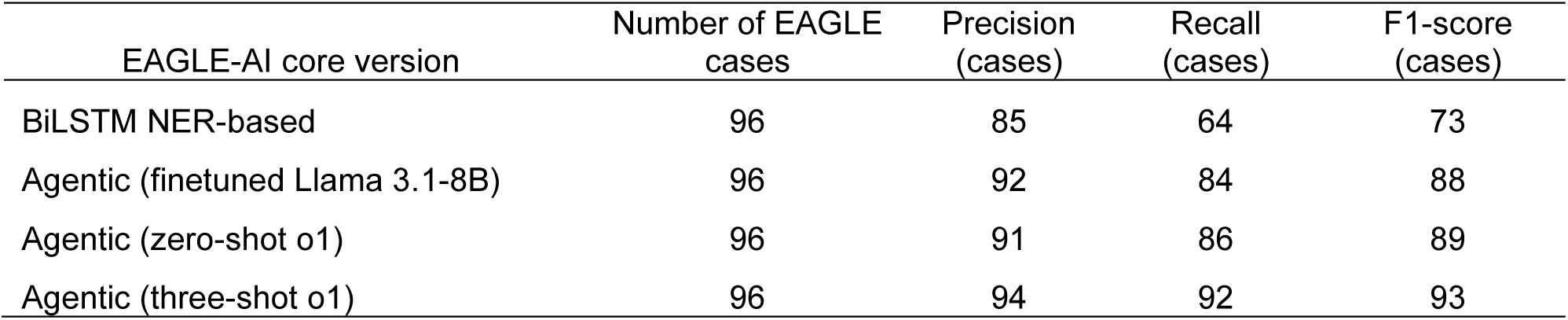
EAGLE-AI performance, ASD case extraction.

**Table S6:** EAGLE-AI performance, genomic variant ID extraction.

| EAGLE-AI core version | Number of EAGLE cases | Precision (variants) | Recall (variants) | F1-score (variants) |
| --- | --- | --- | --- | --- |
| BiLSTM NER-based | 96 | 78 | 56 | 65 |
| Agentic (finetuned Llama 3.1-8B) | 96 | 88 | 81 | 84 |
| Agentic (zero-shot o1) | 96 | 85 | 80 | 82 |
| Agentic (three-shot o1) | 96 | 90 | 88 | 89 |

**Table S7:** EAGLE-AI performance, extracting phenotype descriptions.

| EAGLE-AI core | Number of EAGLE cases | Precision (phenotype) | Recall (phenotype) | F1-score (phenotype) |
| --- | --- | --- | --- | --- |
| BiLSTM NER-based | 96 | 79 | 62 | 70 |
| Agentic (finetuned Llama 3.1-8B) | 96 | 91 | 83 | 87 |
| Agentic (zero-shot o1) | 96 | 89 | 89 | 89 |
| Agentic (three-shot o1) | 96 | 92 | 90 | 91 |

EAGLE-AI's annotation extraction precision-recall metrics out of 100 using four different approaches, evaluated across all manually scored cases from 116 publications; here, the evaluation set was restricted to cases with a final EAGLE score and with a reported variant in *DMD*, *SHANK1*, *CACNA1D*, *DDX3X*, *DDX53*, or *MECP2*.

**Table S8:** Llama 3.1-8B base v. finetuned response.

|  |
| --- |
| Prompt |
| ""Skewed X-chromosome Inactivation in Women with Idiopathic Intellectual Disability is Indicative of Pathogenic Variants<br>Abstract<br>Intellectual disability (ID) is an early onset impairment in cognitive functioning and adaptive behavior, affecting approximately 1% of the population worldwide. Extreme skewing of X-chromosome inactivation (XCI) can be associated with ID phenotypes caused by pathogenic variants in the X chromosome. We analyzed the XCI pattern in blood samples of 194 women with idiopathic ID, using the androgen receptor gene (AR) methylation assay. Among the 136 patients who were informative, 11 (8%) presented with extreme or total XCI skewing, which was significantly higher than expected by chance. Whole-exome data obtained from these 11 patients revealed the presence of dominant pathogenic variants in eight of them, all sporadic cases, resulting in a molecular diagnostic rate of 73% (8/11 patients). All variants were mapped to ID-related genes with dominant phenotypes: four variants in the X-linked genes DDX3X, WDR45, and PDHA1, and four variants in the autosomal genes KCNB1, CTNNB1, YY1, and ANKRD11.<br>In four patients, the variants occurred in the X-linked genes DDX3X, PDHA1, and WDR45; in four other cases, mutations were disclosed in autosomal genes (YY1, ANKRD11, CTNNB1, and KCNB1).<br>Patient P1: Clinical Features: NPMD, ID, shyness; Family History: None; OMIM Disease: Intellectual developmental disorder, X-linked, syndromic, Snijders Blok type (OMIM#300958); Inheritance: XLD/R; Variant: NM_001356.5:c.1171-2A>G (NP_001347.3:p.?); Gene: DDX3X; Inactivation Status: 100:0""<br>"From the research article, state mutations in the DDX3X gene and their clinical significance." |
| Llama 3.1-8B base response |
| Based on my analysis of the research article, I have identified the mutation NM_001356.5:c.1171-2A>G in the DDX3X gene. However, I'm unable to determine its clinical significance from the available information. The article mentions this variant but doesn't provide conclusive evidence about whether it's pathogenic, benign, or of uncertain significance. Additional information would be needed to assess its impact on protein function, its prevalence in affected populations, and its association with specific clinical phenotypes. To make a proper clinical assessment of this DDX3X variant, more comprehensive genetic and functional studies would be required. |
| Llama 3.1-8B finetuned response |
| Based on the research article, I identified one mutation in the DDX3X gene: Mutation: NM_001356.5:c.1171-2A>G (NP_001347.3:p.?) Clinical significance: This is a pathogenic variant associated with intellectual developmental disorder, X-linked, syndromic, Snijders Blok type (OMIM#300958). The patient (P1) presented with neurodevelopmental disorder (NPMD), intellectual disability (ID), and shyness. The variant shows X-linked dominant/recessive inheritance pattern (XLD/R) and demonstrates complete X-chromosome inactivation skewing (100:0), strongly supporting its pathogenicity. |

Prompt was assembled from Chaves et al. 2022<sup>69</sup> title, abstract, paper text, and supplementary table.

**Table S9:**
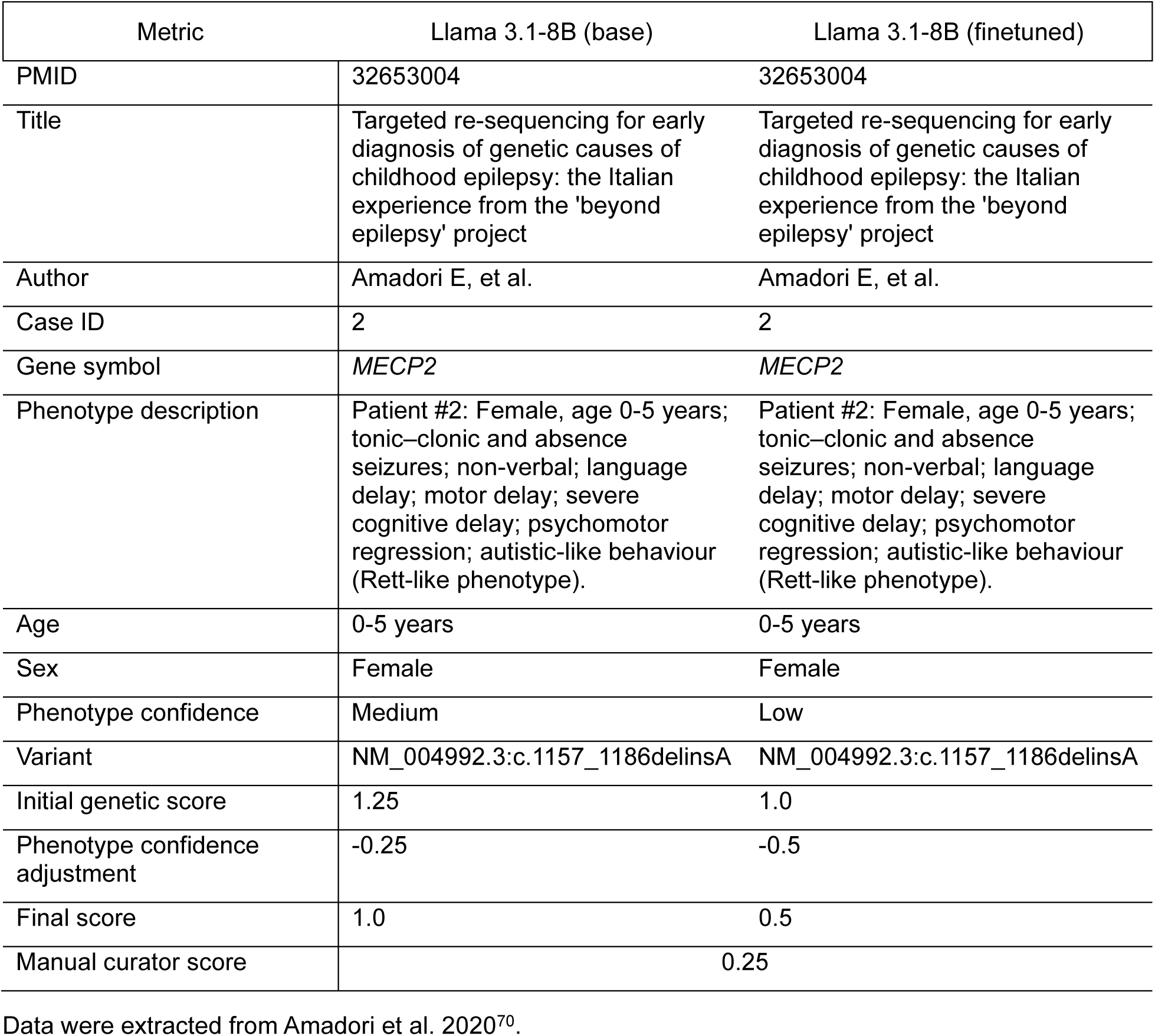
Llama 3.1-8B base v. finetuned data extractions.

**Table S10:** Agentic EAGLE-AI core’s scoring planner agent output instructions.

|  |  |
| --- | --- |
| Required data |  |
| • gene identifier<br>• variant type, inheritance pattern<br>• variant population frequency<br>• ASD diagnostic criteria used, severity of phenotype<br>• ASD phenotype confidence rating<br>• intellectual ability status<br>• experimental evidence (i.e. functional, functional alteration, and rescue)<br>• contradictory evidence<br>• total evidence summary: limited, moderate, strong, definitive |  |
| Output format for each required datum |  |
| datum | "Give name of datum" |
| status | "Give as 'KNOWN', 'NEEDS_SEARCH', 'UNSEARCHABLE', or 'REQUIRES_INPUT'" |
| evidence | "Give supporting evidence or explanation" |
| known_information | "List information already in database" |
| search_topics | "List information that needs additional research" |
| required_inputs | "List information that needs direct input from paper authors" |

Table representation of output instructions contained within the prompt to agentic EAGLE-AI core's scoring planner agent. Instructions are formatted as a Python dictionary within the prompt f-string.

**Table S11:** Agentic EAGLE-AI core’s scoring reporter agent output instructions.

| Section header | Contents |
| --- | --- |
| Gene information | gene identifier, basic information |
| Genetic evidence | variant details<br>variant inheritance pattern<br>population frequency<br>total genetic evidence score (0 – 5.5 per variant) |
| ASD phenotype evaluation | diagnostic / phenotyping methods<br>intellectual ability assessment and cautionary comments, if needed<br>total ASD phenotype evaluation score (high, medium, low) and corresponding adjustment |
| Experimental evidence | functional experiments (score: 0 – 2)<br>functional alteration experiments (score: 0 – 2)<br>model organisms / cell cultures (score: 0 – 4)<br>rescue experiments (score: 0 – 4)<br>total experimental evidence score (0 – 6) |
| Contradictory evidence | disputed or refuting evidence<br>impact on final classification |

**Table S12:** Agentic EAGLE-AI core scoring judge agent output instructions.

|  |  |
| --- | --- |
| overall_assessment | "Give as 'PASS' or 'NEEDS_REVISION'" |
| overall_score | "Give score out of 40" |
| genetic_evidence | score "Give quality score out of 10" |
|  | findings "List quality-related findings for report" |
|  | issues "List quality-related problems or gaps in report" |
| asd_phenotype_evaluation | score "Give quality score out of 10" |
|  | findings "List quality-related findings for report" |
|  | issues "List quality-related problems or gaps in report" |
| experimental_evidence | score "Give quality score out of 10" |
|  | findings "List quality-related findings for report" |
|  | issues "List quality-related problems or gaps in report" |

**Table S13:** Data extraction prompt for deterministic scoring accuracy evaluation.

|  |  |
| --- | --- |
| **1. Identify Relevant Cases** |  |
| Scan the full text (main body, tables, figures, supplements) for individual subjects/patients who meet both of the following:<br>• reported with a genetic variant in the gene of interest<br>• reported with an assessment, diagnosis, or phenotypic features related to ASD |  |
| **2. Extract Key Information** |  |
| Case Identifier | Unique label (e.g. Patient 1, F1-II:1). If absent, describe location (e.g. "paragraph 3"). |
| Case Description | Full description as stated. [Include Quote] |
| Genetic Variant Information | • Gene Name: Record the gene symbol for the case.<br>• Variant Details: HGVS format or description; include coordinates. [Include Quote]<br>• Variant Type: Classify and rate predicted effect (e.g. null, other)<br>• Inheritance Pattern: *de novo*, inherited, unknown. [Include Quote]<br>• Zygosity: Heterozygous, homozygous, hemizygous, etc.<br>• Functional Data: Variant-specific functional evidence. [Include Quote] |
| ASD Phenotype Information | • Diagnosis Method: Tools or descriptions used. [Include Quote]<br>• Phenotype Confidence (A–F): Based on best-supported criterion: A-C = High (expert dx, validated tools, DSM/ICD mentioned), D = Medium (clear symptoms in both ASD domains), E-F = Low (vague terms, single-domain, screeners only) |
| Intellectual Ability (IA) | • Reported IA Level: e.g. IQ, "severe ID", "normal cognition". [Include Quote]<br>• EAGLE IA Category (a–d): a = No/mild/moderate ID b = Severe ID c = Profound ID d = Insufficient info |
| Family/Cohort Information | Note if from SSC, SPARK, ASC, etc. Record affected relatives with phenotype and variant info. |
| Source within Paper | Pinpoint location: e.g. "Table 1, Patient 3", "Page 6", "Supp Fig S2". |
| Focus | Follow EAGLE definitions precisely, esp. for phenotype and IA confidence. Include exact quotes. Do not infer. Mark absent info clearly. |
| Output Format | Detailed, complete extraction. Use exact text from paper. Extract all relevant cases. Do not paraphrase. |

Table representation of data extraction prompt used in NER-based EAGLE-AI core. The prompt is delivered to GPT-4o after the NER model performs relevance filtering for assembling text chunks.

**Table S14:**
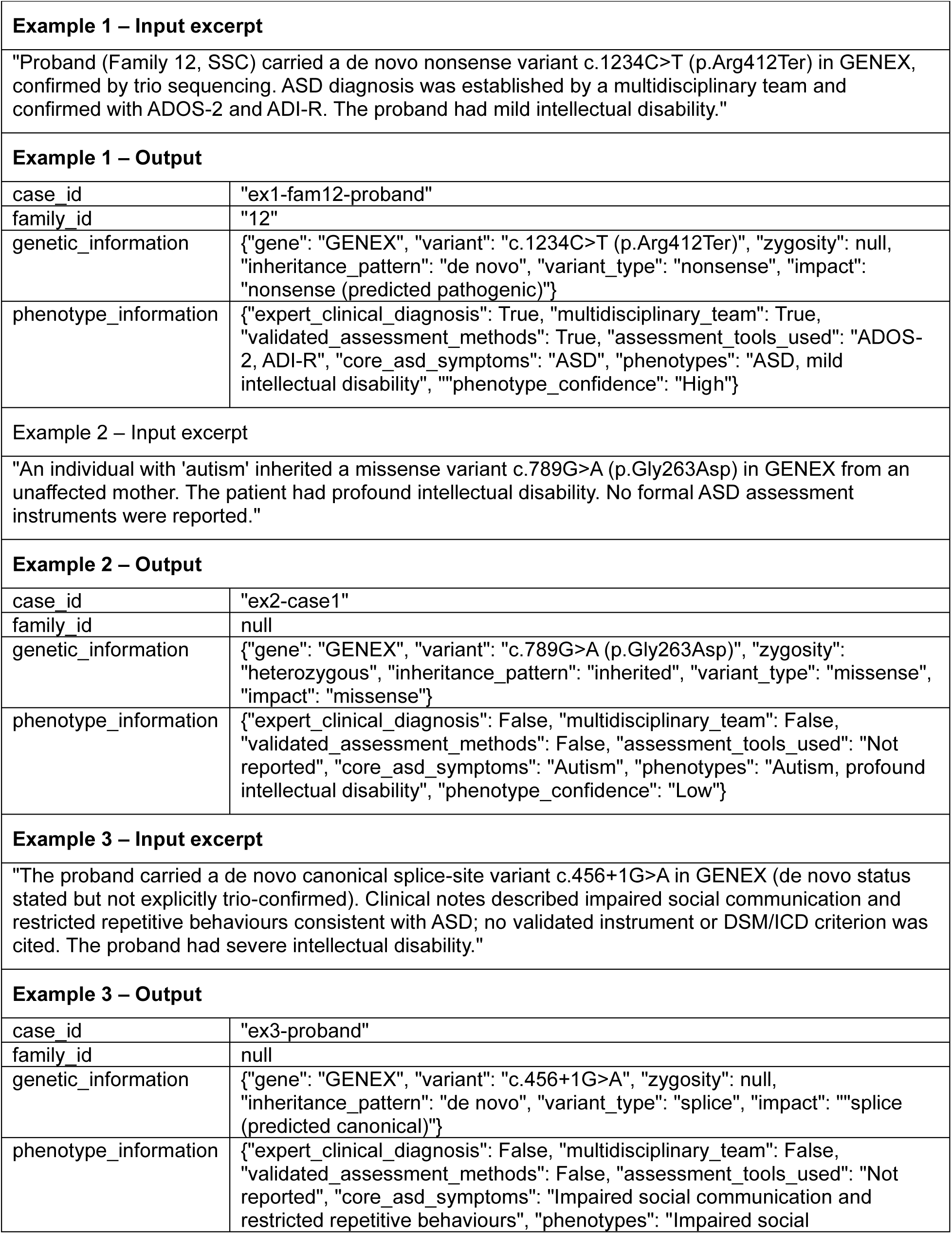
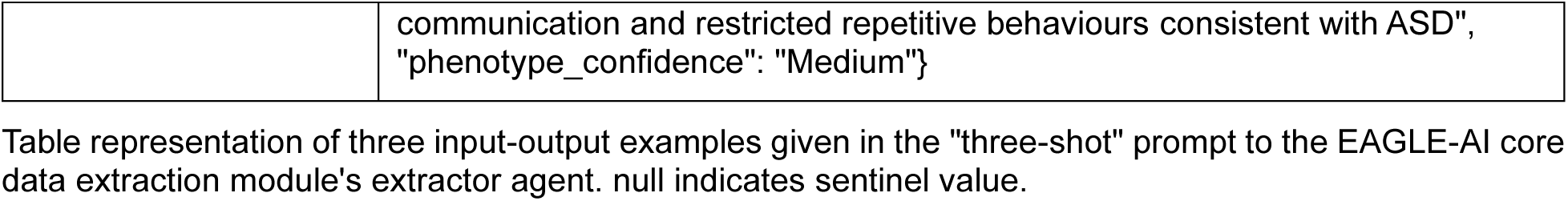
Three input-output examples given in prompt to extractor agent.

**Table S15:** Three input-output examples given in prompt to scoring reporter agent.

| **Example 1 – Input [De novo null, high-confidence phenotype]** |  |
| --- | --- |
| case_id | "P001" |
| family_id | "F001" |
| variant_information | {"variant_type": "nonsense", "impact": "nonsense (predicted pathogenic)",<br>"inheritance_pattern": "de novo"} |
| phenotype_confidence | "High" |
| **Example 1 – Output** |  |
| score_base | 2 |
| phenotype_adjustment | 0 |
| score_final | 2 |
| score_rationale | ""De novo predicted null, no mention of splice → default 2 (0-2.5).<br>No double-count → 2.<br>High confidence → 0 adjustment.<br>Final = 2."" |
| **Example 2 – Input [De novo missense, low-confidence phenotype]** |  |
| case_id | "P002" |
| family_id | "F002" |
| variant_information | {"variant_type": "missense", "impact": "missense (predicted pathogenic)",<br>"inheritance_pattern": "de novo"} |
| phenotype_confidence | "Low" |
| **Example 2 – Output** |  |
| score_base | 0.5 |
| phenotype_adjustment | -0.25 |
| score_final | 0.25 |
| score_rationale | ""De novo missense, no mention of splice → default 0.5 (0-1).<br>No double count → 0.5.<br>Low confidence → -0.25 adjustment.<br>Final = 0.25."" |
| **Example 3 – Input [splicing, sibling de-duplication]** |  |
| case_id | "P003" |
| family_id | "F003" |
| variant_information | {"variant_type": "splice", "impact": "splice (predicted canonical)",<br>"inheritance_pattern": "inherited"} |
| phenotype_confidence | "High" |
| case_id | "P004" |
| family_id | "F003" |
| variant_information | {"variant_type": "splice", "impact": "splice (predicted canonical)", "inheritance_pattern": "inherited"} |
| phenotype_confidence | "High" |
| **Example 2 – Output** |  |
| score_base | 1.5 |
| phenotype_adjustment | 0 |
| score_final | 1.5 |
| score_rationale | ""Inherited canonical splice → default 1.5 (0-1.5).<br>No double count → 1.5.<br>High confidence → 0 adjustment.<br>Final = 1.5."" |
| score_base | 0 |
| phenotype_adjustment | 0 |
| score_final | 0 |
| score_rationale | "" Inherited canonical splice → default 1.5 (0-1.5).<br>From same family → 0.<br>High confidence → 0 adjustment.<br>Final = 0."" |

Table representation of three input-output examples given in the "three-shot" prompt to the EAGLE-AI core scoring module's reporter agent.

**Table S16:**
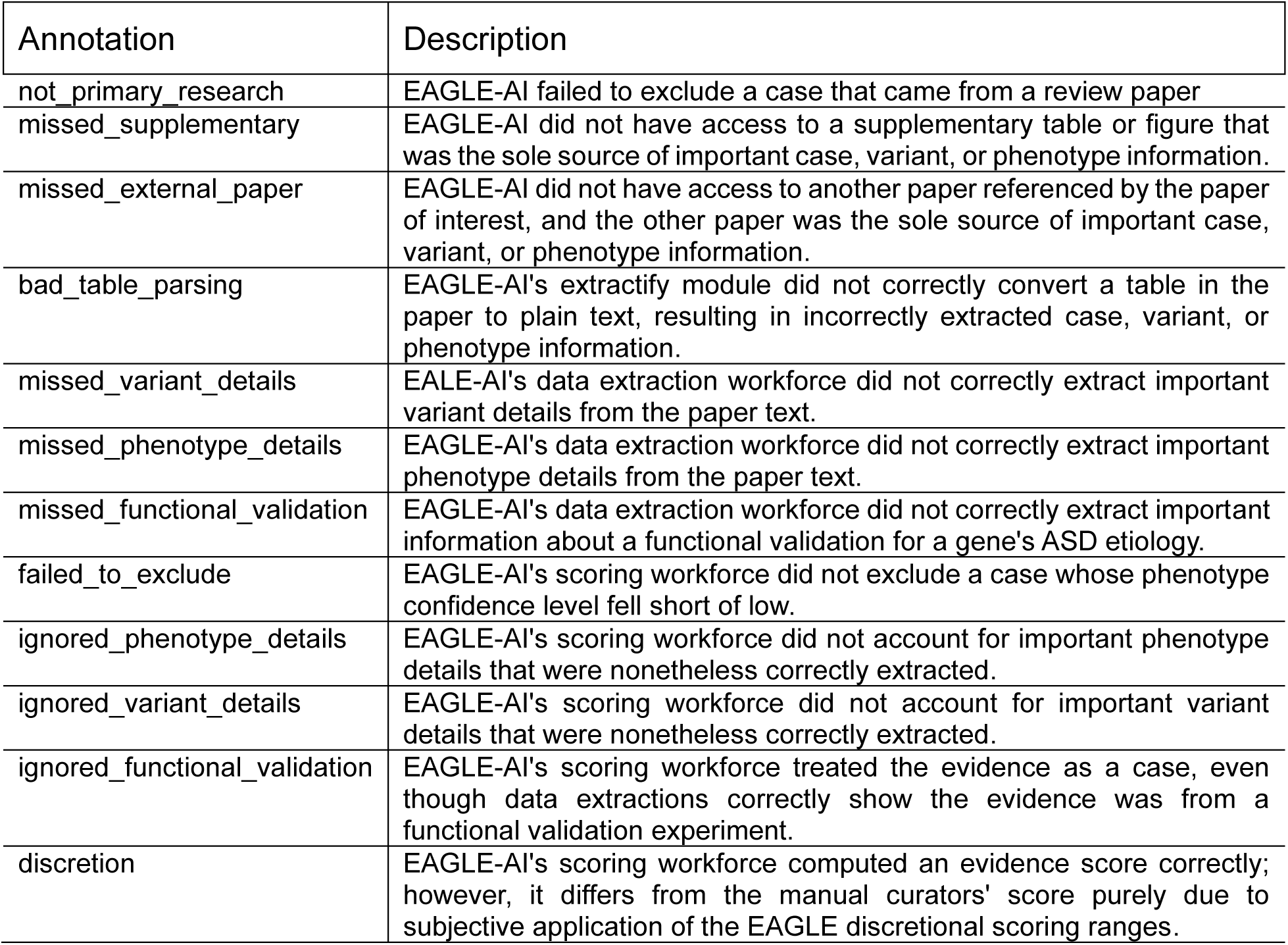
Failure mode annotation descriptions.

| Annotation | Description |
| --- | --- |
| not_primary_research | EAGLE-AI failed to exclude a case that came from a review paper |
| missed_supplementary | EAGLE-AI did not have access to a supplementary table or figure that was the sole source of important case, variant, or phenotype information. |
| missed_external_paper | EAGLE-AI did not have access to another paper referenced by the paper of interest, and the other paper was the sole source of important case, variant, or phenotype information. |
| bad_table_parsing | EAGLE-AI's extractify module did not correctly convert a table in the paper to plain text, resulting in incorrectly extracted case, variant, or phenotype information. |
| missed_variant_details | EAGLE-AI's data extraction workforce did not correctly extract important variant details from the paper text. |
| missed_phenotype_details | EAGLE-AI's data extraction workforce did not correctly extract important phenotype details from the paper text. |
| missed_functional_validation | EAGLE-AI's data extraction workforce did not correctly extract important information about a functional validation for a gene's ASD etiology. |
| failed_to_exclude | EAGLE-AI's scoring workforce did not exclude a case whose phenotype confidence level fell short of low. |
| ignored_phenotype_details | EAGLE-AI's scoring workforce did not account for important phenotype details that were nonetheless correctly extracted. |
| ignored_variant_details | EAGLE-AI's scoring workforce did not account for important variant details that were nonetheless correctly extracted. |
| ignored_functional_validation | EAGLE-AI's scoring workforce treated the evidence as a case, even though data extractions correctly show the evidence was from a functional validation experiment. |
| discretion | EAGLE-AI's scoring workforce computed an evidence score correctly; however, it differs from the manual curators' score purely due to subjective application of the EAGLE discretionary scoring ranges. |

